# The genetic and phenotypic correlates of neonatal Complement Component 3 and 4 protein concentrations with a focus on psychiatric and autoimmune disorders

**DOI:** 10.1101/2022.11.09.22281216

**Authors:** Nis Borbye-Lorenzen, Zhihong Zhu, Esben Agerbo, Clara Albiñana, Michael E. Benros, Beilei Bian, Anders D Børglum, Cynthia M. Bulik, Jean-Christophe Philippe Goldtsche Debost, Jakob Grove, David M. Hougaard, Allan F McRae, Ole Mors, Preben Bo Mortensen, Katherine L. Musliner, Merete Nordentoft, Liselotte V. Petersen, Florian Privé, Julia Sidorenko, Kristin Skogstrand, Thomas Werge, Naomi R Wray, Bjarni J. Vilhjálmsson, John J. McGrath

**Author notes:** equal first author (Nis Borbye-Lorenzen, Zhihong Zhu).

## Abstract

The complement system, including complement components 3 and 4 (C3, C4), traditionally has been linked to innate immunity. More recently, complement components have also been implicated in brain development and the risk of schizophrenia. Based on a large, population-based case-cohort study, we measured the blood concentrations of C3 and C4 in 68,768 neonates. We found a strong correlation between the concentrations of C3 and C4 (phenotypic correlation = 0.65, *P*-value < 1.0×10^−100^, genetic correlation = 0.38, *P*-value = 1.9×10^−35^). A genome-wide association study (GWAS) for C4 protein concentration identified 36 independent loci, 30 of which were in or near the major histocompatibility complex on chromosome 6 (which includes the *C4* gene), while six loci were found on six other chromosomes. A GWAS for C3 identified 15 independent loci, seven of which were located in the *C3* gene on chromosome 19, and eight loci on five other chromosomes. We found no association between (a) measured neonatal C3 and C4 concentrations, imputed C4 haplotypes, or predicted *C4* gene expression, with (b) schizophrenia (SCZ), bipolar disorder (BIP), depression (DEP), autism spectrum disorder, attention deficit hyperactivity disorder or anorexia nervosa diagnosed in later life. Mendelian randomisation (MR) suggested a small positive association between higher C4 protein concentration and an increased risk of SCZ, BIP, and DEP, but these findings did not persist in more stringent analyses. Evidence from MR supported causal relationships between C4 concentration and several autoimmune disorders: systemic lupus erythematosus (SLE, OR and 95% confidence interval, 0.37, 0.34 – 0.42); type-1 diabetes (T1D, 0.54, 0.50 - 0.58); multiple sclerosis (MS, 0.68, 0.63 - 0.74); rheumatoid arthritis (0.85, 0.80 - 0.91); and Crohn’s disease (1.26, 1.19 - 1.34). A phenome-wide association study (PheWAS) in UK Biobank confirmed that the genetic correlates of C4 concentration were associated a range of autoimmune disorders including coeliac disease, thyrotoxicosis, hypothyroidism, T1D, sarcoidosis, psoriasis, SLE and ankylosing spondylitis. We found no evidence of associations between C3 versus mental or autoimmune disorders based on either MR or PheWAS. In general, our results do not support the hypothesis that C4 is causally associated with the risk of SCZ (nor several other mental disorders). We provide new evidence to support the hypothesis that higher C4 concentration is associated with lower risks of autoimmune disorders.

## Introduction

The complement systems are an integral part of the innate immune response^1-4^. These phylogenetically-ancient systems involve complex and interlinked amplification cascades, which can be triggered to protect the body from pathogens. Elements of the system are also involved in a range of additional physiological functions. For example, a growing body of evidence links elements of the complement systems (e.g. Complement Component 4; C4) to brain development and psychiatric disorders^5-7^.

Several of the genes that encode components of the complement system, including *C4*, are located within the major histocompatibility complex (MHC). Linking disease phenotypes with loci within the MHC is difficult because of the long-range linkage disequilibrium (LD) in this region. Furthermore, the *C4* gene has two homologous isoforms (*C4A* and *C4B*), each of which can vary according to an insertion of a human endogenous retrovirus (*HERV*) transposon, and which can vary between one to three genocopies per haplotype^8^. Sekar and colleagues^9^ proposed that genetic variants involving the *C4* gene could account for the strong signal over this region detected in genome-wide association studies (GWAS) of schizophrenia^10^. They found that the expression of mRNA transcripts coding for the *C4A*-related isoform was increased in post-mortem schizophrenia brain samples (cases = 35, controls = 70), and also reported that the *C4A* copy number was associated with both increased *C4A* expression in the brain and increased risk of schizophrenia. These findings are of interest to neurodevelopmental disorders, given evidence that *C4* and related members of the complement systems are involved in synaptic pruning during early brain development^9,11-13^. Apart from the links with schizophrenia, increased *C4A* copy number has also been associated with a *decreased* risk of autoimmune disorders^14,15^. In light of the shared genetic architecture between different types of mental disorders^16^, and the links between *C4A* alleles and risk of autoimmune disorders, there is a need to explore if the putative risk haplotypes are associated with a wider range of both mental- and autoimmune disorders.

As more copies of the *C4A* gene are associated with increased expression of *C4A-*related transcripts in the brain^9,17^, it is reasonable to assume that an increased copy number of the *C4A* gene would also be reflected in increased expression of the C4 protein. C4 is an abundant circulating protein, produced mainly in the liver, and evidence from transgenic mouse experiments^12^ and human observational studies^18,19^ confirms a dose-response relationship between increased *C4* copy number and increased concentration of C4 protein in the peripheral circulation. The concentration of the C4 protein is correlated with other members of the complement family, including complement component C3 (encoded by the *C3* gene)^20^. A recent systematic review and meta-analysis found that the serum concentrations of C4 and C3 did not differ in those with schizophrenia versus controls^21^. However, the neurodevelopmental hypothesis of schizophrenia suggests that early life disruption of brain development may underpin the subsequent adult-onset disorder^22,23^. Additional evidence suggests that complement-related synaptic pruning may be most prominent during early life (e.g. C3 protein expression peaks in the early post-natal period and decreases with age)^7,24^. We are aware of one study that examined neonatal C4 concentration in a schizophrenia case-control study (cases = 75, controls = 644)^25^. This study found evidence that an increased concentration of one of the two measured peptides within the protein encoded by *C4A* was associated with an increased risk of subsequent schizophrenia. There is a need to explore the association between complement-related protein concentrations and later mental disorders based on larger samples of neonatal blood samples.

We examined the association between neonatal C3 and C4 concentrations and later health outcomes based on a population-based case-cohort study that had access to archived neonatal dried blood spots^26,27^. Building on the only published GWAS of serum complement components C3 and C4 (studies based on 3,495 Han Chinese men^18^), we completed GWASs for C3 and C4 neonatal concentrations based on 68,768 samples and estimated the heritability of these phenotypes. We were also able to assess the association between imputed *C4* haplotypes and observed neonatal C4 protein concentration. We examined the association of both (a) imputed *C4* haplotypes and (b) observed C3 and C4 protein concentration in neonatal dried blood spots versus the risk of a range of clinically observed mental disorders in the case-cohort study (i.e. schizophrenia, bipolar disorder, depression, autism spectrum disorder, attention deficit hyperactivity disorder and anorexia nervosa). Based on the results of the GWASs of C3 and C4, we used: (a) bioinformatics tools to explore gene properties of the genome-wide significant loci (summary-data-based MR [SMR], gene-based analyses and gene set analyses); (b) Mendelian randomization analyses to explore the associations between C3 and C4 protein concentrations versus a range of mental disorders (as listed above) and autoimmune disorders (multiple sclerosis [MS], type-1 diabetes [T1D], Crohn’s disease [CD], ulcerative colitis [UC], rheumatoid arthritis [RA], and systemic lupus erythematosus [SLE]), and finally, (c) phenome-wide association studies (PheWAS)^28^ to examine the relationship between the genetic correlates of C3 and C4 protein concentrations versus mental and autoimmune disorders, as well as a wide range of other health phenotypes. A summary of the overall methods is shown in Figure 1.

**Figure 1.**
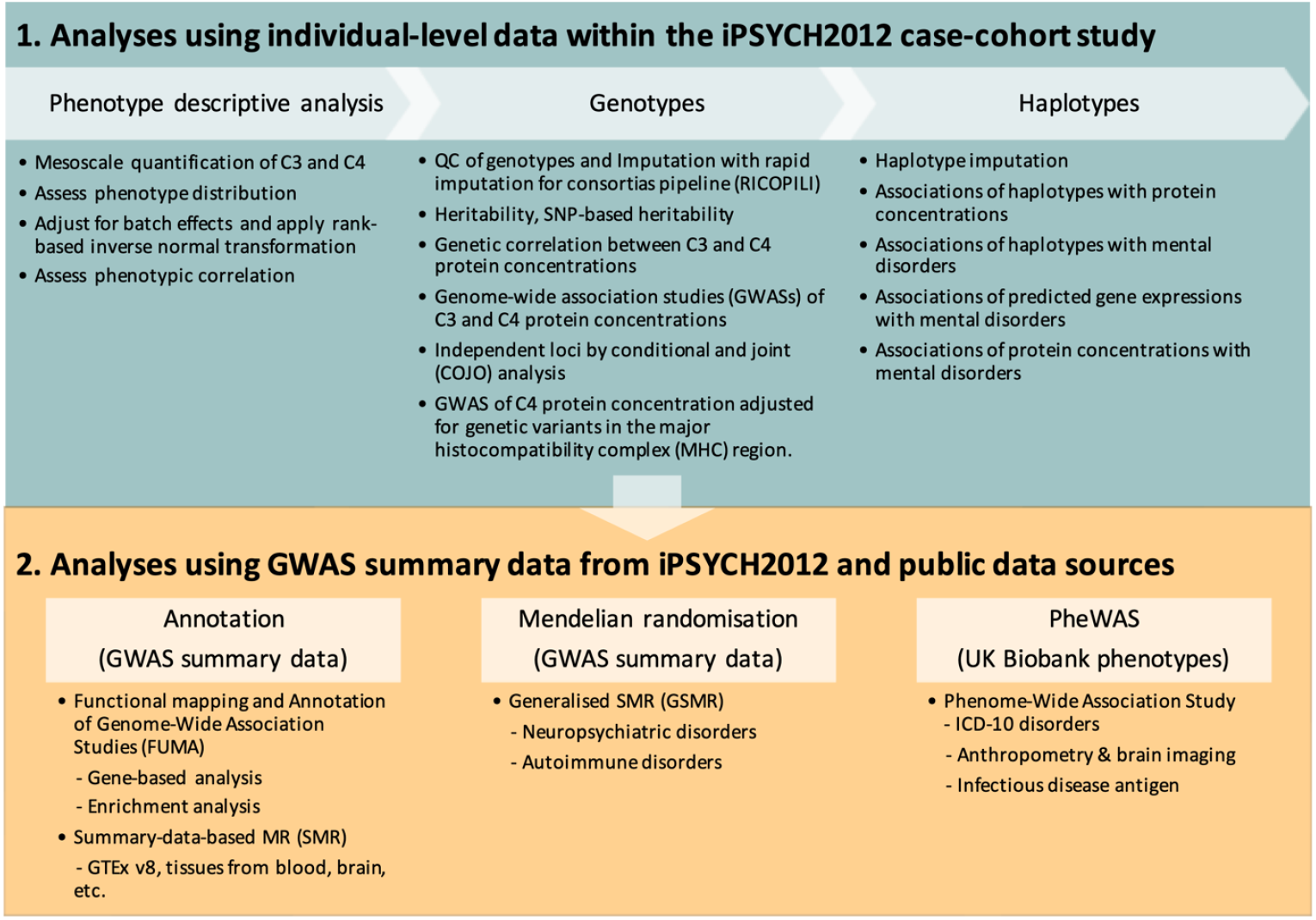
Methods figure.

## METHODS

### The iPSYCH2012 study

Key elements of this study were based on the Lundbeck Foundation Initiative for Integrative Psychiatric Research (iPSYCH) sample^26^, a population-based case-cohort designed to study the genetic and environmental factors of schizophrenia (SCZ), bipolar disorder (BIP), depression (DEP), autism spectrum disorder (ASD) and attention deficit hyperactivity disorder (ADHD). The original iPSYCH sample (known as iPSYCH2012) included information on case status complete through 31 December 2012. We also included 4,791 anorexia nervosa cases (AN; ANGI-DK) from the Anorexia Nervosa Genetics Initiative (ANGI)^29^, which had the same design as iPSYCH2012. Henceforth, we refer to iPSYCH2012 as the combined dataset with the ANGI samples. The iPSYCH2012 sample is nested within the entire Danish population born between 1981 and 2005 (*n*=1,472,762). Diagnoses were identified in the Danish Central Psychiatric Research Register^30,31^, which includes all inpatient contacts in Danish psychiatric hospitals since 1969 and all outpatient and emergency contacts since 1995. The ICD-10 codes used to classify the psychiatric disorder cases can be found in **Supplementary Table 1**. The phenotype information for the iPSYCH2012 participants was updated for the target mental disorders until December 2016. The case-cohort sample includes a population-based random sub-cohort^32^ (*N* = 30,000) with an inclusion probability of 2.04% of the study base (30,000 / 1,472,762). This sub-cohort also includes some participants with the target mental disorders of interest. The genotypes and C3 and C4 protein concentrations were measured in neonatal dried bloodspots (DBSs) taken as part of routine screening at birth from all babies born in Denmark since 1981 and stored in the Danish Neonatal Screening Biobank^33^. Dried blood spot samples have been collected from practically all neonates born in Denmark since 1st May 1981 and stored at −20 °C. Samples are collected 4–7 days after birth. After the dried blood spots were retrieved from the biobank, samples were extracted in a PBS buffer and stored for further use at - 80°C. Subsequently, DNA was extracted according to previously published methods^34^. After storage the protein extracts were assayed for C3 and C4 concentrations. Thus, all genotypes and C3/C4 protein concentration data originated from a single DBS extraction. Additional details related to blood spot extraction and storage are provided in **Supplementary Methods 1**.

### Ethical framework

Material from the Danish Neonatal Screening Biobank has been used primarily for screening for congenital disorders, but are also stored for follow-up diagnostics, screening, quality control and research. According to Danish legislation, material from The Danish Neonatal Screening Biobank can be used for research after approval from the Biobank, and the relevant Scientific Ethical Committee. There is also a mechanism in place ensuring that one can opt out of having the stored material used for research. The Danish Data Protection Agency and the Danish Health Data Authority approved this study. According to Danish law, informed consent is not required for register-based studies. All data accessed were deidentified.

### C3 and C4 protein concentrations

These methods have been described in a related study^35^. Two 3.2 mm discs of DBS were punched into 96 well polymerase chain reaction plates (72.1981.202, Sarstedt). The extracts were analyzed with a multiplex immunoassay (also measuring vitamin D binding protein^35^) using U-plex plates (Meso-Scale Diagnostics (MSD), Maryland, US) employing antibodies specific for complement C3 (HYB030-07 and HYB030-06) and complement C4 (MA1-72520 (ThermoFisher Scientific) and HYB162-04). The antibodies were purchased from SSI Antibodies (Copenhagen, Denmark) except if otherwise stated. Extracts were analyzed diluted 1:70 in diluent 101 (#R51AD, MSD). Capture antibodies (used at 10 μg/mL as input concentration) were biotinylated in-house using EZ-Link Sulfo-NHS-LC-Biotin (#21327, Thermo Fisher Scientific) and detection antibodies were SULFO-tagged (R91AO, MSD), both at a challenge ratio of 20:1. As calibrators, we used complement components purified from human: C3: #PSP-109 (Nordic Biosite, Copenhagen, DK), C4: abx060108 (Abbexa, Cambridge, UK). Calibrators were diluted in diluent 101, detection antibodies (used at 1 μg/mL) were diluted in diluent 3 (#R50AP, MSD). Controls were made in-house from part of the calibrator solution in one batch, aliquoted in portions for each plate, and stored at -20°C until use. The samples were prepared on the plates as recommended by the manufacturer and were read on the QuickPlex SQ 120 (MSD) 4 min after adding 2x Read buffer T (#R92TC, MSD). Analyte concentrations were calculated from the calibrator curves on each plate using 4PL logistic regression using the MSD Workbench software.

Intra-assay variations were calculated from 38 measurements analyzed on the same plate of a pool of extract made from 304 samples. Inter-assay variations were calculated from controls analyzed in duplicate on each plate during the sample analysis, 1022 plates in total. Lower limits of detections were calculated as 2.5 standard deviations from 40 replicate measurements of the zero calibrator. The higher detection limit was defined as the highest calibrator concentration. The lower and upper detection limits for: (a) C3 were 95.4 and 7.98×10^4^ μg/L respectively, and (b) C4 were 55.2 and 7.98×10^4^ μg/L respectively. The intra- and inter-assay coefficient of variation (CV) for (a) C3 were 5.2% and 18.1% respectively; and for (b) C4 were 3.9% and 8.5% respectively. To validate the stability of the samples during storage, we randomly selected 15-16 samples from five years (1984, 1992, 2000, 2008, and 2016; a total of 76 samples). After extracting the samples and adding them to an MSD plate, the rest of the extracts were frozen for 2 months, thawed and measured as described above to imitate the freeze-thaw cycle of the samples in the study. The oldest samples (from 1984) recorded higher concentrations, most probably due to a change in the type of filter paper after 1989. In light of this artefact, we adjusted all DBP values by plate (the sequence of testing followed the date of birth of the sample). This is described below. Additional details related to pre-analytic variation are provided in **Supplementary Methods 1**.

### Imputation of genotypes

DNA genotyping was conducted at the Broad Institute (Boston, MA, USA) using the Infinium PsychChip v1.0 array (Illumina, San Diego, CA, USA)^36^. We restricted the genotyped SNPs to 252,339 high quality, common SNPs based on build hg19 (the same human genome reference build was used throughout this study). Details of the filtering can be found elsewhere (Schork et al., 2019). Briefly, we excluded SNPs with minor allele frequency (MAF) < 0.01, Hardy Weinberg Equilibrium (HWE) *p*-value < 1.0×10^−6^ or non-SNP alleles (i.e., insertions and deletions, INDELs). 245,328 autosomal and 7,011 X-chromosome (chrX) SNPs were retained and used to impute SNPs using the Ricopili pipeline^37^ with the Haplotype Reference Consortium (HRC)^38^ as the imputation reference panel (accession number: EGAD00001002729). 6,743,499 autosomal SNPs, 227,371 chrX SNPs for males and 184,517 chrX SNPs for females were retained with missing rate < 0.02 and genotype call probability > 0.8. We further excluded the imputed SNPs with imputation info score < 0.8, MAF < 0.01 or HWE *p*-value < 1.0×10^−6^. 5,201,724 SNPs were retained in autosomes and 126,109 SNPs were retained on chrX. We then used the common SNPs to infer the genetic ancestries of 80,873 participants in the iPSYCH2012 study, 75,764 individuals of European ancestry and 5,109 individuals of non-European ancestry. Details are provided in **Supplementary Method 2**.

### Imputation of C4 haplotypes

*C4* haplotypes were imputed from reference data^9,14^ using the genotyped SNPs in the iPSYCH2012 sample. The human *C4* haplotypes have various copy numbers, including two isotypic polymorphisms, *C4A* (*A*) and *C4B* (*B*). Each isotype has two length-polymorphisms due to a human endogenous retroviral (*HERV*) insertion, long form (*L*, with *HERV* insertion) and short form (*S*, without *HERV* insertion). The isotypic and length polymorphisms lead to four alleles in a *C4* copy, *AL, AS, BL* and *BS*. Using the genotyped SNPs, the *C4* haplotype reference was used to impute the C4 alleles and the number of *C4* copies (with a maximum copy number of 4). The *C4* haplotype imputation panel comprised whole genome sequencing data from 1,234 individuals of multiple ancestries, which enabled us to identify *C4* alleles with high accuracy. We used Beagle software^39^ for the imputation with the *C4* haplotype reference. The imputation results provided the counts of alleles, but were unable to confidently distinguish all combinations of variants, for example, between the haplotypes *AS-BL* and *AL-BS*. We counted the two *C4* alleles (*C4A* and *C4B*) with combination of *HERV* using a subset of the imputed result, where combinations can be confidently distinguished (details are provided in **Supplementary Method 3**). Both counts of *C4* allele combinations and reported studies^40^ indicated that the *C4A* gene is more likely to carry *HERV* insertion than the *C4B* gene. Therefore, the *C4* haplotype is assumed to be *AL-BS* rather than *AS*-*BL*, consistent with methods described by Sekar et al^9^. The imputed counts were converted to the *C4* haplotypes. Eight common *C4* haplotypes (allele frequencies ≥ 0.01) were imputed in the iPSYCH2012 study (**Supplementary Table 3**). The allele frequencies of the 8 haplotypes were consistent with other studies^14,41^. Given the common *C4* haplotypes, we counted the copy numbers of the *C4* alleles (**Supplementary Figure 2**) for each participant. 28 individuals (0.04%) carried 4 copies of *C4B* and 35 individuals (0.05%) carried 6 copies of *HERV* insertion. Therefore, we excluded these individuals with very rare copy numbers. The *C4A* copy number is strongly correlated with *C4B* and *HERV* copy numbers (Pearson correlation between *C4A* and *C4B* = -0.52; between *C4A* and *HERV* = 0.73).

### Quality control of the C3 and C4 protein concentrations

The C3 and C4 protein concentrations were measured in 78,268 iPSYCH2012 participants of multiple ancestries. We focused on 68,768 individuals of European ancestry with measures of C3 and C4 protein concentrations. The protein assay plates captured a substantial amount of variance (C3 = 49.4%, C4 = 45.3%). Therefore, we used a linear mixed model (LMM)^42^ approach to adjust protein concentrations, *y* = ∑*z*_plate_*u*_plate_ + *e*, where *y* represents the C3/4 protein concentration; *z*_plate_ represents protein assay plate, a random variable; *u*_plate_ represents the random effect of protein assay plate; and *e* represents residual. The mixed model regression was conducted by the R package of lme4 (Bates et al., 2015). The rank-based inverse normal transformation (RINT)^43^ was applied to the residuals to have mean 0 and variance 1. The standard deviations (SDs) adjusted for variance captured by protein assay plate were used for the interpretation of results of C3 and C4 protein concentrations, for C3 protein concentration, 1 SD unit = 2.56 μg/L (3.60 μg/L × √(1 - 0.49)), and for C4 protein concentration, 1 SD unit = 2.46 μg/L (3.33 μg/L × √(1 - 0.45)).

### Heritability and SNP-based heritability of the C3 and C4 protein concentrations

The iPSYCH2012 cohort had 75,764 participants of European ancestry. 19,113 participants who shared a genetic relatedness (entry of genetic relationship matrix, *r*_GRM_) ≥ 0.05 with at least one other individual were considered as relatives; 3,253 first degree (*r*_GRM_ ≥ 0.4), 2,077 second degree relatives (0.2 ≤ *r*_GRM_ < 0.4) and 13,783 third degree relatives (0.05 ≤ *r*_GRM_ < 0.2). We jointly estimated both the heritability (*h*^2^) and the SNP-based *h*^2^ (*h*^2^_SNP_) of the C3 and C4 protein concentrations by using the method proposed by Zaitlen et al^44^. This method assumes a normal distribution of SNP effect sizes. The GWAS studies of protein concentrations^45-47^ observed that SNPs in or near the coding genes would capture more phenotypic variance than the remaining SNPs. Therefore, we used two approaches to further explore the *h*^2^_SNP_, 1) estimating it using all common SNPs by BayesR^48^, which assumes a mixture distribution of SNP effect sizes and can be used to test number of SNPs with nil, small, median and large genetic variance (small *R*^2^ < 0.01%, median 0.01% ≤ *R*^2^ < 0.1%, and large 0.1% ≤ *R*^2^ < 1%) in addition to estimation of *h*^2^_SNP_, and 2) partitioning *h*^2^_SNP_ into (a) *h*^2^_cis-chr_, explained by SNPs on the chromosome where the coding gene (cis-chr SNPs) was positioned, and (b) *h*^2^_trans-chr_, explained by the remaining SNPs (trans-chr SNPs). To partition *h*^2^_SNP_, we divided SNPs into two subsets, 1) cis-chr and 2) trans-chr SNPs. The *h*^2^_cis-chr_ and *h*^2^_trans-chr_ were estimated by using GCTA-GREML^49^ and BayesR. Only genetically unrelated participants were included in the two analyses. The genetic relationship matrix used in the Zaitlen and GREML analyses were estimated from 5,201,724 common SNPs. Only the subset of HapMap phase 3 (HM3) SNPs were included in the BayesR analyses because of the computation complexity (853,129 HM3 SNP in total; for C3 protein concentration, 132,239 cis-chr SNPs and 839,891 trans-chr SNPs; for C4 protein concentration, 60,631 cis-SNPs and 792,498 trans-SNPs). The Zaitlen method and GREML were implemented in Genome-wide Complex Trait Analysis (GCTA)^50^. BayesR was implemented in Genome-wide Complex Trait Bayesian analysis (GCTB). The URLs for these programs are provided below.

We estimated the genetic correlation between C3 and C4 concentrations by BOLT-REML^51^. To further examine if the genetic correlation was primarily driven by the two protein-coding genes (i.e. *C3* and *C4*), we conducted the BOLT-REML and Haseman-Elston regression^52^ analyses using the trans-chr SNPs. The Haseman-Elston regression was implemented in GCTA.

### GWAS of C3 and C4 protein concentrations

We performed the GWAS analysis of the C3 and C4 protein concentrations by fastGWA^53^. The fastGWA is a LMM method which can include all individuals of European ancestry regardless of relatedness. 5,201,724 imputed SNPs were analysed in the GWAS. In addition, fastGWA can include candidate markers which optimizes power if particular SNPs capture a large proportion of the total variance^54^. Therefore, we excluded the SNPs in and near the coding gene for the required GRM in the GWAS, C3: chr19, 4.67Mb – 8.74Mb, C4: chr6, 24.8Mb – 33.9Mb. For each GWAS, we fitted birthyear, sex, wave (i.e., genotyping batch) and the first 20 PCs as covariates in the model. The PCs were estimated by FastPCA^55^, excluding the same SNPs as we did for the required GRM. We conducted the GWASs using all SNPs on autosomal and sex chromosomes. SNPs on X chromosome for males (coded as 0/2) were tested as diploid, assuming X chromosome of males has half dosage compensation^56^. We used GCTA-COJO^50^ to identify the SNPs which were independently associated with the two concentrations. We randomly sampled 10,000 participants from the population-based sub-cohort of iPSYCH2012 as the LD reference cohort. The GWAS significance threshold was 5.0×10^−8^.

To explore if the enrichment of mental disorder cases in the iPSYCH2012 case-cohort could induce bias within the GWASs, we conducted simulations with ascertained individuals and performed GWASs in the population-based sub-cohort (**Supplementary Method 4**).

### Associations between C4 haplotypes and protein concentrations

We examined the associations between the imputed *C4* haplotypes and the two observed C3 and C4 protein concentrations. We first examined the associations of *C4* copy numbers using a LMM approach, *y*_protein_ = *x*_copy_*b*_copy_ + ∑*x*_*c*_*b*_*c*_ + ∑*z*_-MHC_*u*_-MHC_ + *e*, where *y*_protein_ was C3/4 protein concentration; *x*_copy_ was copy number of *C4* allele, either *C4A, C4B* or *HERV*; *b*_copy_ represents effect of copy number; *x*_*c*_ was covariate with *b*_*c*_ being its effect; Both *b*_copy_ and *b*_*c*_ were fixed effects. The covariates in the model were the same as those fitted in the GWAS of C4 protein concentration. Because of the strong linkage disequilibrium in the MHC region, fitting SNPs in the region is likely to underestimate the effects of *C4* allele count and reduce power. Therefore, we fitted the SNPs outside the MHC region (*z*_-MHC_) in the model with *u*_-MHC_ being their random effects. In practice, the effect of copy number from the linear mixed model could be estimated by generalized least squares (GLS) method, *b* = (X^*T*^V^-1^X)^-1^X^*T*^V^-1^*y*_protein_, where X = {*x*_copy_, *x*_*c*_} and V was phenotypic covariance matrix of C3/4 protein concentration. It was implemented by GCTA-GREML. All the individuals of European ancestry were included in the analysis. The three *C4* allele counts were correlated. Therefore, we estimated the joint effects using the same LMM approach as above, *y*_protein_ = *x*_C4A_*b*_C4A_ + *x*_C4B_*b*_C4B_ + *x*_HERV_*b*_HERV_ + ∑*x*_*c*_*b*_*c*_ + ∑*z*_-MHC_*u*_-MHC_ + *e*. In the model, *x*_C4A_, *x*_C4B_ and *x*_HERV_ represent respectively counts of *C4A, C4B* and *HERV*. The remaining variables were defined as above. Secondly, we further examined the associations of the imputed *C4* haplotypes using the LMM approach. Previous studies have reported a strong effect for the *C4A* gene^9^, while the effect of *C4B* remains unclear^57^. Therefore, we used ‘*BS*’ as the reference haplotype to estimate joint effects of the remaining haplotypes. The regression model can be expressed as, *y*_protein_ = ∑*x*_allele_*b*_allele_ + ∑*x*_*c*_*b*_*c*_ + ∑*z*_-MHC_*u*_-MHC_ + *e*, where *x*_allele_ represents *C4* haplotype. Seven *C4* haplotypes were included in the model, except for BS. The remaining parameters were defined as above. The estimated effect can be interpreted as the effect of the *C4* haplotype compared to BS. All the European participants were included in the analysis. The significance threshold for these analyses was the same as the main GWAS significance threshold (i.e., 5.0×10^−8^).

### FUMA and SMR

We conducted gene-based analysis by Functional Mapping and Annotation of Genome-Wide Association Studies (FUMA)^58^. There were 18,305 genes available for the gene-based analysis, thus the Bonferroni corrected threshold was 1.4×10^−6^ (= 0.05 / (18,305 × 2)). We conducted Summary-data-based Mendelian Randomisation (SMR)^59^ to identify pleiotropic genes for C3 and C4 concentrations. For the SMR analysis, the eQTL data (i.e., summary statistics from associations of gene expressions), was Genotype-Tissue Expression version 8 (GTEx v8)^60^. The LD reference sample with 10,000 participants was the same as the above GCTA-COJO analysis. 22,338 gene-tagged probes within 49 tissues (200,144 probes in total) which had significant SNPs were included in the SMR analysis. The Bonferroni significance threshold was 1.2×10^−6^ (= 0.05 / (200,144 × 2)).

### Associations between C4 haplotypes and mental disorders observed within the iPSYCH2012 case-cohort study

Based on the associations with protein concentrations, we conducted the associations between *C4* haplotypes and 6 iPSYCH2012 disorders (SCZ, BIP, DEP, ASD, ADHD and AN). We used three approaches to examine the relationships, 1) associations with *C4* allele counts, 2) associations with imputed *C4* haplotypes, 3) associations with predicted *C4* gene expression in the brain. Because the iPSYCH2012 case-cohort study has person-level data on the age-at-first contact with psychiatric services, we were able to assess the risk of mental disorders within the more informative time-to-event framework, using Cox proportional hazards regression (Cox PH) to analyse the hazards of *C4* allele counts and haplotypes with respect to the mental disorder of interest. For *C4* allele count, we examined the joint effects due to their correlations, *h*(*t*) = *h*_0_(*t*)exp(*x*_C4A_*b*_C4A_ + *x*_C4B_*b*_C4B_ + *x*_HERV_*b*_HERV_ + ∑*x*_*c*_*b*_*c*_). In the model, *h*_0_(*t*) represents the baseline hazard while *h*(*t*) represents the hazard at time *t* between baseline and December 2016. The remaining variables were defined as above. For C4 haplotypes, we examined the joint effects using the Cox PH model, *h*(*t*) = *h*_0_(*t*)exp(∑*x*_allele_*b*_allele_ + ∑*x*_*c*_*b*_*c*_). All the variables were defined as above. In addition to C4 haplotypes, we used the predicted *C4A* and *C4B* gene expressions as outlined in the post-mortem brain study of Sekar et al.^9^ The association was conducted with a Cox PH model, *h*(*t*) = *h*_0_(*t*)exp(*x*_C4A_predicted_*b*_C4A_predicted_ + *x*_C4B_predicted_ *b*_C4B_predicted_ + ∑*x*_*c*_*b*_*c*_), where *x*_C4A_predicted_ and *x*_C4B_predicted_ represent the predicted *C4A* and *C4B* gene expressions, respectively. We included only unrelated individuals of European ancestry in the three analyses.

In the time-to-event analysis, the cases were the diagnosed participants by December 2016, and the non-cases are defined as the entire cohort excluding those individuals with the disorder of interest. Therefore, we defined six psychiatric-disorder samples for the time-to-event analyses. The sample sizes for cases and non-cases are shown in **Supplementary Table 1**.

### Associations between protein concentrations and mental disorders observed within the iPSYCH2012 case-cohort study

Based on the associations between *C4* haplotypes and 1) the two protein concentrations (C3 and C4) and 2) six mental disorders, we explored the associations between C3 and C4 protein concentrations and mental disorders observed in the iPSYCH2012 case-cohort study, using Cox PH models. We first tested the marginal effects of two concentrations, using the model *h*(*t*) = *h*_0_(*t*)exp(*x*_protein_*b*_protein_ + ∑*x*_*c*_*b*_*c*_). The variables were defined as above. Due to the high correlation, we then fitted both concentrations jointly, *h*(*t*) = *h*_0_(*t*)exp(*x*_C3_protein_*b*_*C3_*protein_ + *x*_C4_protein_*b*_*C4_*protein_ + ∑*x*_*c*_*b*_*c*_), where *x*_C3_protein_ and *x*_C4_protein_ represent C3 and C4 concentration, respectively. *b*_*C3_*protein_ and *b*_*C4_*protein_, effects of two protein concentrations, were fixed effects. In the three analyses, we included only unrelated individuals of European ancestry.

### Mendelian Randomisation analysis based on summary statistics

We explored the relationships between protein concentrations and mental and autoimmune disorders using the generalised summary-data-based Mendelian Randomisation (GSMR) method^61^. Because of the possible link between C3 and C4 and brain function^7^, we also included two neurodegenerative disorders—Alzheimer’s disease, and amyotrophic lateral sclerosis in these analyses. Thus, there were 8 broadly-defined neuropsychiatric disorders (i.e., SCZ^62^, DEP^63^, BIP^64^, ASD^65^, ADHD^66^, AN^67^, Alzheimer’s disease^68^, amyotrophic lateral sclerosis^69^), and 6 autoimmune disorders (i.e., multiple sclerosis^70^, type-1 diabetes^71^, Crohn’s disease^72^, ulcerative colitis^72^, rheumatoid arthritis^73^, systemic lupus erythematous^74^). The GWAS summary statistics for these disorders were publicly available (additional details provided in **Supplementary Table 19)**.

Unfortunately, detailed GWAS summary statistics for Sjögren’s syndrome were not available. The GSMR method was implemented in GCTA. The GSMR method includes options to exclude potentially pleotropic loci (HEIDI filtering). Similar to the methods for pleiotropy exclusion, it assumes fewer pleiotropic SNPs than valid variants and MR estimates may fluctuate with the inclusion of those SNP outliers^61^. Given that a substantial proportion of the variance in C4 concentration was associated with SNPs in the MHC region (including the *C4* gene), genetic variants used in the GSMR analysis may be dominated by those positioned in or near the region. Therefore, the interpretation of results based on the large effect loci within the MHC can be misleading. As a planned analyses, we ran the GSMR method again based on summary statistics from the GWAS of C4 concentration adjusted for COJO SNPs within MHC region (as a covariate). In addition, in the presence of bidirectional GSMR findings (i.e., evidence of both protein concentration impacting on phenotype of interest, and vice versa; forward and reverse direction respectively), symmetrical (i.e., equivalent) effect sizes may reflect the presence of pleiotropy. It is of note that effects of C3/4 protein concentration on these mental and autoimmune disorders by GSMR were equivalent to log odds ratio (OR) from logistic regression if we have all the phenotypes in a cohort^61^. We, therefore, reported the logOR and 95% CI in the results. The effects of mental and autoimmune disorders on C3/4 protein concentration may be required to be transformed for better interpretation^75^. However, we only used the reserve GSMR results to examine the presence of reverse causation and pleiotropy. Therefore, we reported the GSMR estimates directly. The LD reference sample which included 10,000 participants was the same as for the GCTA-COJO analysis. We conducted both forward (from C3/4 concentration to disorder) and reverse (from disorder to C3/4 concentration) analyses. We set HEIDI-outlier threshold at 0.01 to filter horizontal pleiotropy. The GSMR Bonferroni corrected significance threshold was 1.9×10^−3^ (= 0.05 / (2 × 13)).

### PheWAS based on UK Biobank phenotypes

Based on the GSMR analysis results, we conducted phenome-wide association studies (PheWASs) to explore the relationships with disorder outcomes. The PheWASs were conducted in the UK Biobank (UKB) cohort^76^, a large population cohort with 487,409 participants of multiple ancestries. The genotypes were imputed to the HRC^38^ and UK10K^77^ reference panels by the UKB group. The quality controls were described in detail elsewhere^78^, including generic ancestry determination, quality controls of imputed SNPs, and estimation of principal components. In the study, we included 1,130,559 HM3 SNPs on autosomal chromosomes, with MAF ≥ 0.01, HWE *P*-value ≥ 1.0 ×10^−6^, because only effects of HM3 SNPs were predicted by BayesR. The genetic relationship matrix was estimated by GCTA. 347,769 unrelated participants of European ancestry were retained with genetic relationship < 0.05. In the PheWAS analysis, we included 1,148 UKB phenotypes, 1) 1,027 disorders which were classified by ICD-10 codes, 2) 51 anthropometric measurements and brain imaging traits, and 3) 70 infectious disease antigens. The quantitative traits were standardised by RINT to have mean 0 and variance 1. We then used the model to test the associations, for quantitative traits, *y* = *x*_protein_prs_*b*_protein_prs_ + ∑*x*_*c*_*b*_*c*_ + e, where *y* represents quantitative trait in UKB; *x*_protein_prs_ represents polygenic scores for neonatal C3 and C4 protein concentrations predicted by BayesR; *x*_*c*_ represent the covariate variables including birth year, sex and 20 PCs. For dichotomous traits, logit(*y*) = *x*_protein_prs_*b*_protein_prs_ + ∑*x*_*c*_*b*_*c*_ + e, where *y* represents the dichotomous trait and definitions of the remaining variables were the same as above. In addition, we conducted the PheWAS analyses for males and females separately using the same approach. Polygenic scores were predicted using GWASs in both sexes. The Bonferroni corrected significance threshold was 7.3×10^−6^ (= 0.05 / (1148 × 3 × 2)).

## RESULTS

The iPSYCH2012 study, a population-based case-cohort, was designed to study the genetic and environmental factors of 6 mental disorders (**Supplementary Table 1**), SCZ, BIP, DEP, ASD, ADHD and AN. The study included 80,873 individuals of multiple ancestries. 75,764 European individuals were retained by principal components (PCs) projection (**Supplementary Figure 1**). The following analyses in our study were based on the European individuals. We imputed C4 haplotypes using the reference data^9,14^. The imputation process predicted the counts of three *C4* alleles (*C4A, C4B* and *HERV*), but is not able to confidently distinguish all combinations. From the subset of imputation result without ambiguity, *C4A* allele is more likely to carry a *HERV* than *C4B* allele (∼1.5 higher) while *C4A* allele is much less likely to not carry a HERV (**Supplementary Table 2**, for *C4A*, 0.2%, for *C4B*, 21.2%), which is consistent with previous studies^40^. Based on that, eight common *C4* haplotypes were imputed with allele frequency (AF) ≥ 0.01 (**Supplementary Table 3**). Their frequencies were respectively *BS* (12%), *AL* (4%), *AL-BS* (23%), *AL-BL* (43%), *AL-BS-BS* (2%), *AL-AL* (11%), *AL-AL-BS* (3%), and *AL-AL-BL* (2%), consistent with the published studies^14,41^. We counted the copy numbers of the three types of *C4* alleles (*C4A, C4B* and *HERV*, **Supplementary Figure 2**). The copy numbers (i.e., count) for the different types of *C4* alleles were correlated. *C4A* count was negatively correlated with *C4B* count (*r* = -0.52, *P*-value < 1.0×10^−100^). *HERV* count was positively correlated with *C4A* count (*r* = 0.73, *P*-value < 1.0×10^−100^), but negatively correlated with *C4B* count (*r* = -0.17, *P*-value < 1.0×10^−100^).

There were 68,768 participants of European ancestry with measures of C3 and C4 protein concentrations. The distributions of the observed neonatal C3 and C4 protein concentrations were right skewed, with mean, median, SD, and interquartile range being respectively 7.1, 6.7, 3.6, and 5.1 - 9.2 μg/L for C3 protein concentration, and 6.9, 6.5, 3.3 and 4.9 - 9.0 μg/L for C4 protein concentration (**Supplementary Table 4**). Significant differences were observed in the protein concentrations between males and females (C3 protein concentration: difference = -0.32, SE = 0.03, *P*-value = 6.5×10^−32^; C4 protein concentration: difference = -0.30, SE = 0.03, *P*-value = 2.7×10^−33^).

While the variance captured by sex was small (*R*^2^ = 0.19% for C3 protein concentration and 0.20% for C4 protein concentration), we fitted sex as a covariate in the following analyses. To account for the influence of duration of storage (**Supplementary Figure 3**) and between-protein assay plate variation, we regressed the concentrations of the plates using a linear mixed model (LMM) approach. The residuals were standardised (mean 0, variance 1) using rank-based inverse normal transformation (RINT). After standardisation, the concentrations of C3 and C4 were positively correlated *r*_*P*_ = 0.65 (*P*-value < 1×10^−100^, **Supplementary Figure 4**).

### Heritability of C3 and C4 protein concentrations

The *h*^2^ of C4 by Zaitlen’s method^44^ was 40% (SE = 0.03, *P*-value = 2.7×10^−44^, **Supplementary Table 5**) while the *h*^2^_SNP_ was 26% (SE = 0.006, *P*-value < 1.0×10^−100^). For C3, *h*^2^ was 21% (SE = 0.03, *P*-value = 1.1×10^−11^) and the *h*^2^_SNP_ was 4% (SE = 0.005, *P*-value = 3.2×10^−14^). The high genetic variance of C4 concentration was confirmed by BayesR^48^ — the *h*^2^_SNP_ for C4 was 24% (SE = 0.004, *P*-value 1.0×10^−100^ and 6% (SE = 0.005, *P*-value = 4.4×10^−39^) for C3. Moreover, ∼50 HM3 SNPs captured substantial genetic variance (*R*^2^ > 0.1%), for C3 protein concentration 39 SNPs and for C4 protein concentration 62 SNPs. The observation was consistent with our expectation that SNPs in or near the salient encoding genes would capture a substantial proportion of phenotypic variance for these two circulating proteins. Thus, we partitioned SNPs into two subsets; (a) those on the chromosome where the coding gene is located (*cis*-chr SNPs), and (b) those on the remaining chromosomes (*trans*-chr SNPs), and jointly estimating SNP-based *h*^2^ at the two subsets of SNPs, SNP-based *h*^2^_cis-chr_ and *h*^2^_trans-chr_, using GREML^49^. For C4, *h*^2^_cis-chr_ = 14% (SE = 0.005, *P*-value < 1.0×10^−100^) and *h*^2^_trans-chr_ = 4% (SE = 0.006, *P*-value = 9.4×10^−13^). For C3, *h*^2^_cis-chr_ was 0.4% (SE = 0.001, *P*-value = 1.1×10^−3^) and *h*^2^_trans-chr_ = 3% (SE = 0.006, *P*-value = 1.4×10^−7^). The cis-chr SNPs of C4 concentration captured more genetic variance than the trans-chr SNPs. A higher genetic variance was found at cis-chr SNPs by BayesR. For C4, *h*^2^_cis-chr_ = 22% (SE = 0.003, *P*-value < 1.0×10^−100^) and *h*^2^_trans-chr_ = 5% (SE = 0.004, *P*-value = 6.2×10^−30^). For C3, *h*^2^_cis-chr_ = 2% (SE = 0.001, *P*-value = 3.9×10^−80^) and *h*^2^_trans-chr_ = 5% (SE = 0.004, *P*-value = 5.9×10^−26^). Both the Zaitlen method and GREML assume genetic variance at each SNP follows a normal distribution while BayesR assumes a mixture distribution. The BayesR estimates are expected to better model the true underlying genetic architecture. We only used individual-level-data-based methods (such as Zaitlen’s method, GREML and BayesR) in the analysis as genotypes and phenotypes were all available, since summary-level-data-based methods modelling LD from reference cohorts are less accurate. In summary, both C3 and C4 concentrations were heritable traits. SNPs positioned in the *C4* gene accounted for a substantial proportion of the genetic variance of C4 concentration.

The genetic correlation (*r*_*g*_) between the two concentrations based on BOLT-REML^51^ was 0.38 (**Supplementary Table 6**, SE = 0.03, *P*-value = 1.9×10^−35^), smaller than the phenotypic correlation (0.65). Given the large genetic effects at the coding genes for C3 and C4 we then estimated *r*_g_ using SNPs other than chromosomes 6 or 19 (related to the location of *C4* and *C3* genes, respectively) to further investigate if the correlation was driven by cis-chr SNPs, *r*_*g*_ = 0.82 (SE = 0.05, *P*-value = 4.8×10^−65^). The high correlation was confirmed by Haseman-Elston regression^52^ (*r*_g_ = 0.78, SE = 0.19, *P*-value = 4.1×10^−5^), using trans-chr SNPs. These results indicated that C3 and C4 were genetically correlated, and this genetic correlation was not driven by SNPs in or near their respective encoding genes.

### GWAS of C3 and C4 protein concentrations

We used fastGWA^53^ to conduct the GWAS analysis based on 75,764 participants of European ancestry and 5,327,833 common SNPs, 5,201,724 in autosomes and 126,109 on the X chromosome (**Figure 2**). We conducted a GCTA-COJO^79^ analysis to help identify putative independent SNPs. For C4 protein concentration, 34 autosomal SNPs were identified as genome-wide significant (**Supplementary Table 7)**, and all were autosomal. For C3 protein concentration, 14 significant SNPs were identified, again all were autosomal (**Supplementary Table 8**).

**Figure 2.**
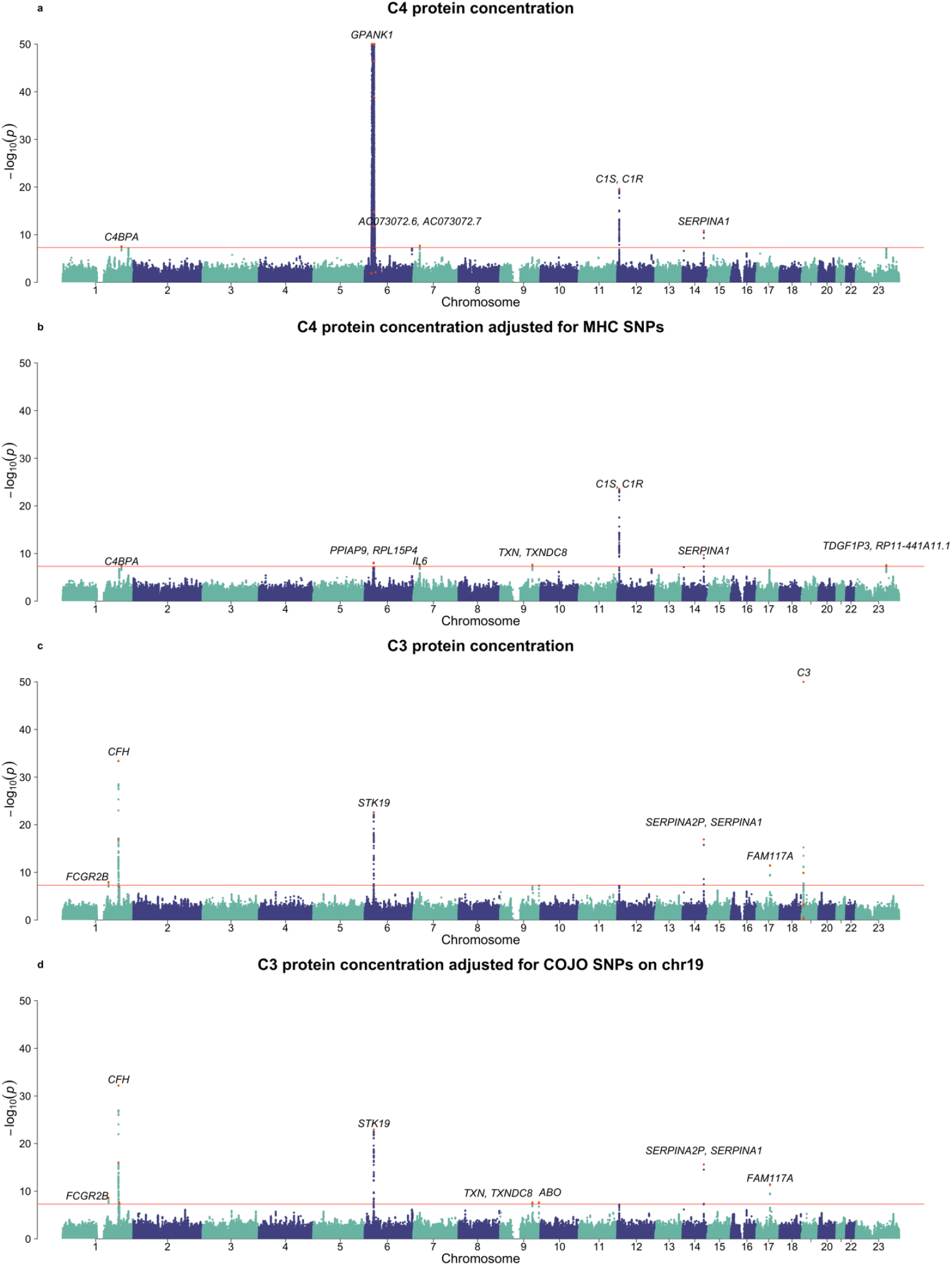
GWASs of neonatal C4 and C3 protein concentrations. a) unadjusted C4 protein concentration, b) C4 protein concentration adjusted for COJO SNPs in the MHC region (fitted as covariates in the regression model), c) unadjusted C3 protein concentration and d) C3 protein concentration adjusted for COJO SNPs on chromosome 19. The COJO SNPs fitted as covariates in GWAS of adjusted protein concentration (panels b and d) were identified from GCTA-COJO analysis of unadjusted protein concentration. The COJO SNPs were highlighted with red colour. The top-associated SNPs were annotated with their overlapped or nearest genes. The GWAS threshold was 5.0×10^−8^.

Of the 34 SNPs significantly associated with C4 protein concentration, 30 (88.2%, 30/34) were found on chromosome 6. Of these 29 were in the MHC region and 27 (79.4%, 27/34) SNPs were positioned within 2Mb of the *C4* gene (chr6, 31.9 Mb). These 27 SNPs explained 16.7% of phenotypic variance in C4 concentration, which is consistent with the estimated *h*^2^_cis-chr_. SNP rs113720465 (32,005,355bp, ∼1Kb away from *C4B-AS1* [32,000-32,004Kb]) had the largest effect size (the A allele was associated with an increase of 0.76 standard deviation units of C4 protein concentration), however SNP rs3117579 had the smallest *P*-value (within an exon of *GPANK1*). Given this large effect size, it is possible that SNPs in LD at *r*^2^ < 0.01 (the COJO threshold of independence) could also be reported as genome-wide significant through correlation. Thus, we conducted a GWAS fitting the COJO SNPs in and near MHC region as fixed effects (**Figure 2**). We identified 8 significant loci by COJO, 6 of which were significant from GWAS of unadjusted C4 protein concentration. The 2 additional loci were on chromosomes 9 (rs6477754) and X (rs12012736). Interestingly, nearly all the 8 COJO SNPs were annotated to the genes biologically related to complement-related pathways (**Supplementary Figure 5**). For example, *C4BPA* (rs12057769) encodes a binding protein of C4. The *IL6* gene (rs2066992) encodes a cytokine stimulated in response to infections and injuries. *C1S* (7.1Mb on chr12) and *C1R* (7.2Mb on chr12), the nearest genes of rs11064501, are the protein-coding genes of two C1 sub-components.

With respect to C3 protein concentration, 7 COJO SNPs were positioned within 2Mb of the *C3* gene (chr19, 6.7Mb) — these loci explained 3% of phenotypic variance in C3 concentration. After fitting these 7 COJO SNPs as covariates, 8 significant COJO SNPs were identified (**Supplementary Figure 6**). We found a SNP within the *ABO* gene, which has recently been identified as a ‘master regulator’ of plasma protein concentration^46,80^. The gene annotations of the remaining SNPs encode proteins which involve immune- and/or C3-related pathways: (a) *FCGR2B* (rs844), which encodes an inhibitory receptor for the Fc region of immunoglobulin gamma (IgG), (b) *CFH* (rs558103 and rs11580821) which encodes Complement Factor H, a key factor that inhibits the alternative pathway and the amplification loop downstream from C3, (c) *STK19* gene (rs114492815) which is close to the *C4A* gene, and (d) *FAM117A* (rs12949906), which has enhanced gene expressions in dendritic cells (i.e., antigen-presenting cells involved in the immune system^81^.

For both GWASs of C3 and C4 protein concentration, we found no evidence of potential ascertainment bias related to the enrichment of cases with mental disorders in the iPSYCH2012 case-cohort study (**Supplementary Method 4, Supplementary Figures 8-10**). Therefore, the following post-GWAS analyses were based on the results from the full iPSYCH2012 sample.

### Associations between C4 haplotypes and C3/4 protein concentration

Both *h*^2^_SNP_ and GWAS results indicated strong effects of the SNPs in the MHC region for both C4 and C3 protein concentrations. Due to the complex LD structure in this region, we used the imputed C4 haplotypes to investigate phenotypic associations of these genetic variants. We first examined the associations between the imputed C4 haplotypes with the observed C4 protein concentration, using an LMM approach. As expected, more copies of *C4* allele (either *C4A, C4B*, with or without *HERV*) were strongly associated with higher C4 protein concentration (**Figure 3**). The *C4A* copy number (*b*_C4A_) had greater effect than *C4B* (*b*_C4B_) and *HERV* (*b*_HERV_), *b*_C4A_ = 0.3 (**Supplementary Table 9**, SE = 0.01, *P*-value < 1.0×10^−100^), *b*_C4B_ = 0.2 (SE = 0.01, *P*-value < 1.0×10^−100^), and *b*_HERV_ = 0.2 (SE = 0.004, *P*-value < 1.0×10^−100^). The *C4* copy numbers were correlated. Therefore, we fitted all 3 gene copy numbers in a regression model to estimate the joint effects. The *C4A* copy number had nearly identical effect to *C4B* copy number, *b*_C4A_ = 0.6 (SE = 0.01, *P*-value < 1.0×10^−100^), *b*_C4B_ = 0.6 (SE = 0.01, *P*-value < 1.0×10^−100^). The beta estimates associated with the *HERV* copy number was less than the comparable estimates for *C4A* and *C4B*, and was negatively associated with C4 protein concentration, *b*_HERV_ = -0.08 (SE = 0.005, *P*-value = 5.0×10^−51^). This may reflect the strong correlation with *C4A* (*r* = 0.73) and negative correlation with *C4B* (*r* = -0.17). The result suggested 1 more copy of *C4A* or *C4B* is likely to have 1.6 μg/L (∼0.6 × SD unit) higher C4 protein concentration given the same amount of *HERV*. We calculated the captured variance (*R*^2^ = *s*^2^*b*^2^) that were comparable between the *C4* copy numbers. In the formula, *s*^2^ was variance of C4 copy number, analogous to variance of allele count. Of interest, the *s*^2^ of *C4A* count was greater than *C4B* count (**Supplementary Table 3**, *s*^2^ = 0.55 for *C4A* and 0.31 for *C4B*). Therefore, *C4A* count had a larger contribution to C4 protein concentration than the *C4B* count (*R*^2^ = 23% for *C4A* and 11% for *C4B*). In total, both counts captured 17.3% of variance in C4 protein concentration, accounting for the negative correlation between the two allele counts (*r* = -0.52). The captured genetic variance was in line with *h*^2^_cis-chr_, and the genetic variance at the MHC SNPs. In summary, the imputed counts of both *C4A* and *C4B* were associated with the observed C4 protein concentration and *C4A* count had a greater contribution than *C4B* count.

**Figure 3.**
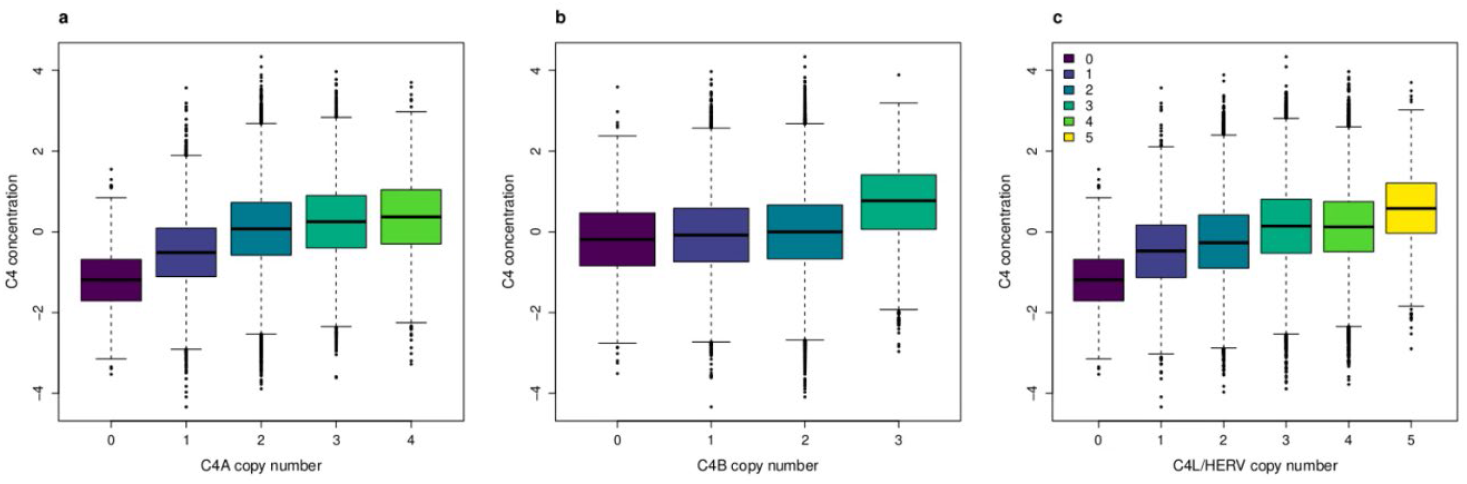
Plot of *C4* copy number versus C4 protein concentration. There were three C4 alleles, (a) *C4A*, (b) *C4B* and (c) *C4L*/*HERV*. The colours represent *C4* allele counts.

Based on the effects of *C4* allele counts, we then examined the association between the commonly observed *C4* haplotypes and C4 protein concentration. For this analysis, we used the *BS* haplotype as the reference category (because of [1] the positive association between *C4* allele count and C4 protein concentration, and [2] the greater contribution of *C4A* count to C4 protein concentration).

All the remaining 7 common haplotypes were associated with increased C4 protein concentration. Due to their higher frequencies, *AL*-*BS* and *AL*-*BL* haplotypes captured greater variance of C4 protein concentration (**Supplementary Table 10**, *R*^2^ = 7.6% for *AL*-*BS* and 7.1% for *AL*-*BL*).

We then examined C3 protein concentration. In keeping with expectations, we did not identify any significant associations between either *C4* copy number or *C4* haplotype, versus C3 protein concentration. However, we found the *AL*-*BS* haplotype was nominally significantly associated with C3 protein concentration (*b*_AL-BS_ = 0.23, SE = 0.03, *P*-value = 5.4×10^−3^). From the GWAS of C3 concentration, there was a COJO SNP positioned within the MHC region (rs114492815). While this SNP was in very weak association with each of the *C4* allele counts (*R*^2^ < 0.005 for *C4A* count and *C4B* count, 0.01 for *C4L*/*HERV* count), there was a moderate association with *AL*-*BS* (*R*^2^ = 0.11). Therefore, we ran the analysis again, fitting rs114492815 as an additional covariate. After this adjustment, none of the haplotypes were associated with the C3 concentration (**Supplementary Table 10**). In general, C3 concentration was independent of C4 alleles.

### Functional mapping of GWAS

Having found the significant SNPs from the GWASs, we explored the genes associated with both concentrations. For C4 concentration, we identified 263 significant genes by Functional Mapping and Annotation of Genome-Wide association Studies (FUMA), 257 (98%) on chromosome 6 and five (2%) on the remaining chromosomes (**Supplementary Table 11**). These findings are consistent with the high LD between loci in the MHC region and the high gene density in this region. Some differentiation between genes was achieved by using SMR which integrates the trait associations with significant eQTL associations. For SMR, using the eQTL summary data GTEx version 8, we identified 56 pleiotropic genes, 55 on chromosome 6 and one on chromosome 1 (**Supplementary Table 12**). We noted that the number of identified genes by SMR was smaller than by FUMA. Many genes on chromosome 6 were significant on the SMR test but failed to pass HEIDI test (i.e., there was evidence of pleiotropy). This was because of the complex LD and likely multiple causal alleles. Interestingly, SMR analysis found that *C4A, C4B* and *C4BPA* were all significantly associated with neonatal C4 protein concentration. These are three of the major genes involved in regulation of C4 protein concentration. The genetic correlates of neonatal C4 protein concentration were associated with higher *C4A* gene expressions in 8 brain tissues, amygdala (*b*_XY_ = 0.70, SE = 0.11, *P*-value = 3.7×10^−10^), anterior cingulate cortex (*b*_XY_ = 0.74, SE = 0.13, *P*-value = 6.8×10^−9^), caudate basal ganglia (*b*_XY_ = 0.70, SE = 0.09, *P*-value = 2.6×10^−16^), cerebella hemisphere (*b*_XY_ = 0.44, SE = 0.05, *P*-value = 8.2×10^−18^), brain cerebellum (*b*_XY_ = 0.43, SE = 0.04, *P*-value = 5.1×10^−22^), hippocampus (*b*_XY_ = 0.68, SE = 0.10, *P*-value = 8.0×10^−12^), hypothalamus (*b*_XY_ = 0.79, SE = 0.11, *P*-value = 4.4×10^−12^), and putamen basal ganglia (*b*_XY_ = 0.76, SE = 0.12, *P*-value = 8.6×10^−10^). Except for cerebellar hemisphere and cerebellum, the effect sizes of these associations were comparable with a mean of *b*_XY_ = 0.73. Overall, the findings from FUMA and SMR indicate strong associations between genes in the MHC region and C4 protein concentration, and the *C4A* gene was likely to have causal effect on C4 protein concentration in brain tissues. Interestingly, these significant genes were enriched with the Kyoto Encyclopedia of Genes and Genomes (KEGG) gene-sets of systemic lupus erythematosus (SLE, *P*-value = 1.1×10^−70^) and complement systems (*P*-value = 7.2×10^−4^) (**Supplementary Figure 7**). For C3 protein concentration, we identified 19 genes by FUMA (**Supplementary Table 13**). Within this set of genes, the *DXO* gene (chr6: 31.9Mb), which is positioned close to rs114492815, a significant SNP from the C3 GWAS, passed both SMR and HEIDI (a test of pleiotropy) tests (**Supplementary Table 14**). All these genes by FUMA and SMR were enriched with KEGG gene-sets of SLE (*P*-value = 3.5×10^−6^), leishmania infection (*P*-value = 5.0×10^−6^), complement systems (*P*-value = 9.1×10^−6^) and Fc gamma R-mediated phagocytosis (*P*-value = 9.5×10^−4^) (**Supplementary Figure 7**). In general, the identified genes (especially those within MHC region) suggest an association between SLE and the complement systems.

### Associations with mental disorders within the iPSYCH2012 case-cohort study

We did not identify significant associations between *C4A, C4B*, or *HERV* copy numbers (**Supplementary Table 15** or C4-related haplotypes (**Supplementary Table 16**) and any of the six mental disorders. Based on the formula between imputed *C4* haplotypes and observed *C4* gene expression (i.e., RNA concentration) in post-mortem brain tissue^9^, we found no significant associations between these estimates and any of the 6 mental disorders (**Supplementary Table 17**). Importantly, we did not find any significant associations between observed neonatal C4 protein concentration and any of the six mental disorders. In models that accounted for the strong correlation between C3 and C4 concentration, we found no significant association between C3 concentration and any of the six mental disorders. (**Supplementary Table 18**).

### GSMR relationships with candidate neuropsychiatric and autoimmune disorders

We conducted Mendelian randomization analyses to examine relationships between the two protein concentrations (C3 and C4) and neuropsychiatric and autoimmune disorders (**Supplementary Table 19**). The results are shown in **Figures 4 and 5**. In the unadjusted analysis (i.e., all loci including the MHC region; with and without HEIDI filtering), higher C4 protein concentration was found to be associated with three mental disorders (**Figure 4 and Supplementary Figure 11**, SCZ, DEP, BIP). The odds ratios for these three findings were small (1.05 or less). We found that these GSMR results were strongly dependent on SNP instruments that were in and near the MHC region (e.g., for schizophrenia and bipolar disorder 126 out of 130 SNPs, and for major depression 103 out of 107 SNPs). These three findings did not persist in the analyses adjusted for the MHC region SNPs. Overall, these analyses do not lend weight to the hypothesis that C4 is causally related to the risk of the psychiatric disorders included in the analyses.

**Figure 4.**
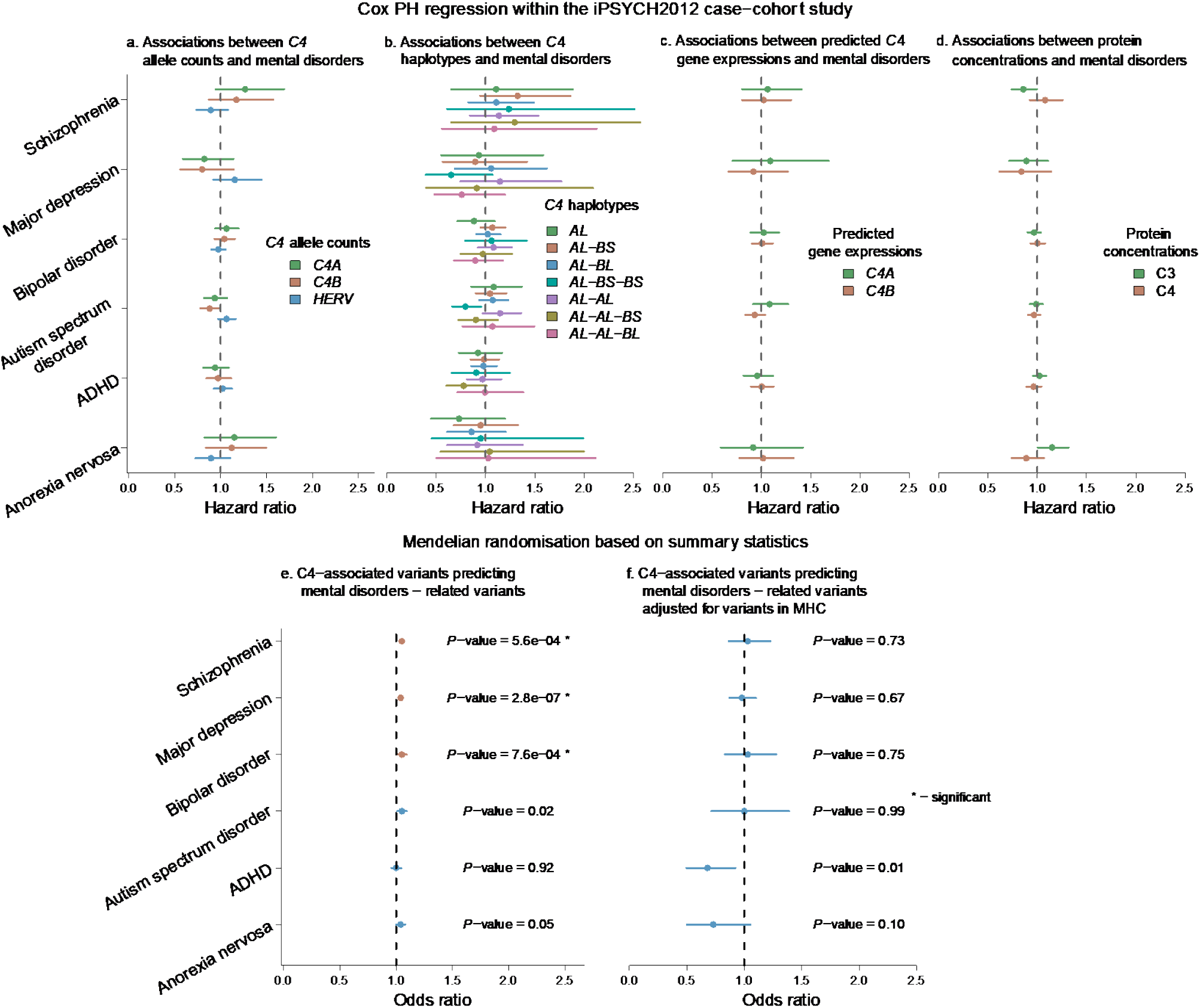
Association between *C4*-related measures and mental disorders, and GSMR analyses with mental disorders. There were 6 mental disorders in the analyses: SCZ, DEP, BIP, ASD, ADHD and AN. The results shown in the top row were from time-to-event analyses between mental disorders and C4-related genotypes and phenotypes, including a) *C4* allele counts, b) imputed *C4* haplotypes, c) predicted *C4* gene expressions and d) C3 and C4 protein concentrations. The analyses were conducted in the iPSYCH2012 cohort. The results from Mendelian randomisation analyses (conducted by GSMR) were shown in the bottom row. The GSMR analysis using GWAS summary statistics predicted relationships between C4 protein concentration and mental disorders, e) using genetic variants from GWAS of C4 protein concentration, f) using genetic variants from GWAS of C4 protein concentration adjusted for COJO SNPs in the MHC region. The Bonferroni corrected thresholds were provided in Methods. Bars represent 95% confidence interval. Significant results were highlighted with “*”.

**Figure 5.**
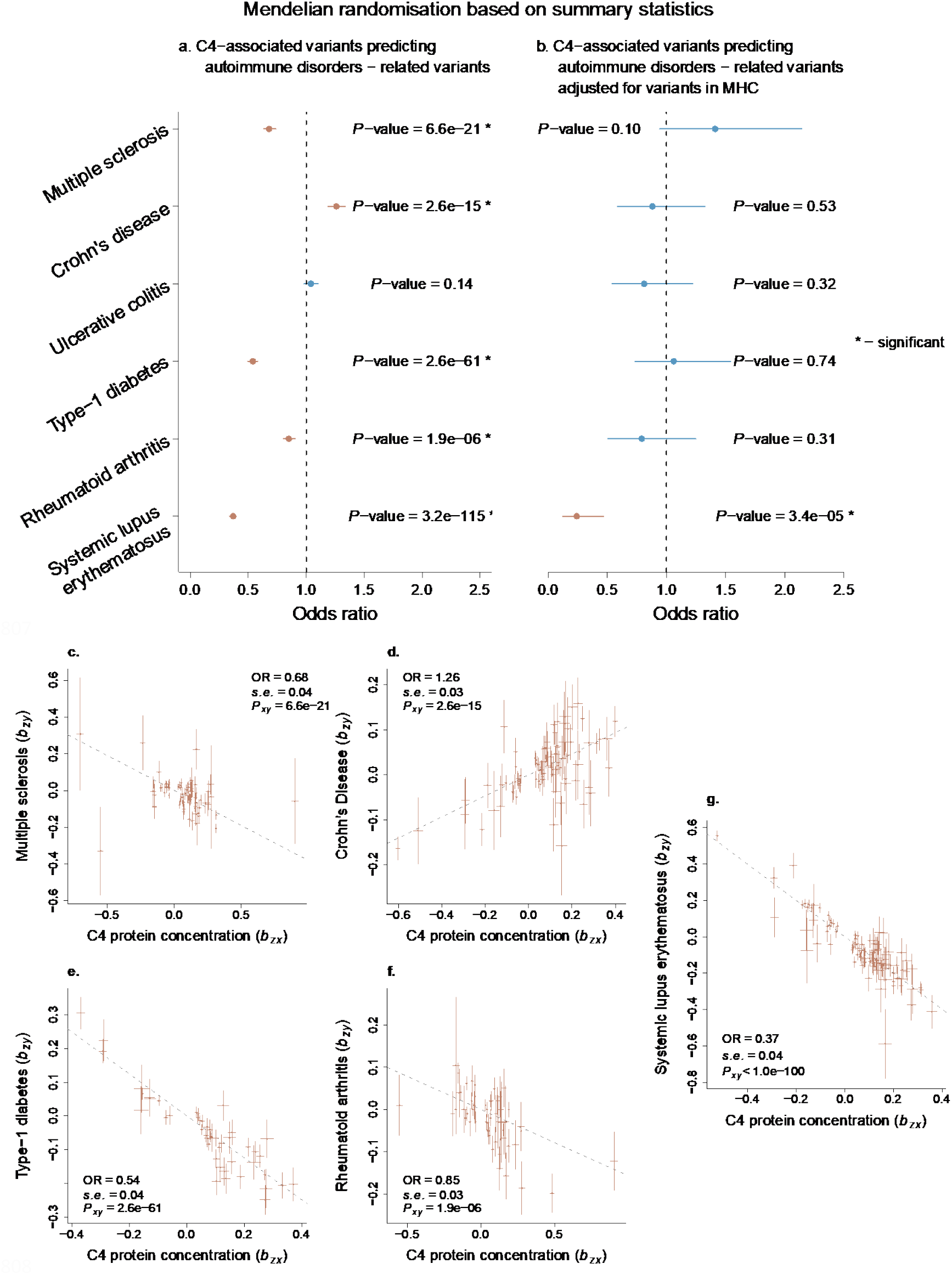
GSMR analyses of C4 protein concentration against autoimmune disorders. The top two panels showed the Mendelian randomisation results a) using genetic variants from GWAS of C4 protein concentration, b) using genetic variants from GWAS of C4 protein concentration adjusted for COJO SNPs in the MHC region. The Bonferroni corrected threshold was 1.8×10^−3^. Bars in the top 2 panels represented 95% confidence interval. Significant results were highlighted with “*”. The lower five panels show the effects of genetic variants for C4 protein concentration without adjustment against effects for five autoimmune disorders. Significant findings were identified for multiple sclerosis, Crohn’s disease, type-1 diabetes, rheumatoid arthritis and systemic lupus erythematosus. The estimates in the panels (OR, standard error and *P*-value) were estimated from GSMR. Slope of dash line represented logOR. Bars in the bottom 5 panels represented standard errors of SNPs from GWAS. Potential pleiotropic SNPs were excluded.

In contrast, we found strong, protective effects of C4 concentration for several autoimmune disorders. Higher C4 concentration was associated with lower risks of multiple sclerosis, type-1 diabetes, rheumatoid arthritis and systemic lupus erythematosus (**Figure 5**). The effects were very large, especially for type-1 diabetes (OR = 0.54, 95% confidence interval [CI] = 0.50 – 0.58, *N*_SNP_ = 47) and systemic lupus erythematosus (OR = 0.37, 95% CI = 0.34 – 0.42, *N*_SNP_ = 103) (**Supplementary Table 20**). We identified that higher C4 concentration increased the risk of Crohn’s disease (OR = 1.26, 95% CI = 1.19 – 1.34, *N*_SNP_ = 86). The strong effect of neonatal C4 protein concentration on these disorders were not caused by reverse causation (**Supplementary Table 21 and Supplementary Figure 12**). After removing the pleiotropic SNPs, genetic variants associated with all autoimmune disorders (except for type-1 diabetes) were not associated with C4 protein concentration in the reverse GSMR analysis (from autoimmune disorder to neonatal C4 protein concentration). When we examined the relationships adjusted for the MHC region SNPs, the significant association with SLE persisted. The effect size was comparable to that found using unadjusted C4 GWAS (with adjustment, OR = 0.24, 95% CI = 0.12 – 0.47, *N*_SNP_ = 7; without adjustment, OR = 0.37, 95% CI = 0.34 – 0.42, *N*_SNP_ = 103). Overall, these findings further support the hypothesis that higher C4 protein concentration is causally related to a reduced risk of systemic lupus erythematosus—it is predicted that an increase of 2.46 μg/L (1 SD unit) of C4 concentration would be associated with a 76% reduced risk (1 – 0.24) of systemic lupus erythematosus.

We then explored the relationships between C3 concentration and neuropsychiatric and autoimmune disorders by bidirectional GSMR. Mindful that analyses based on fewer instruments may be underpowered to detect small effects, no significant associations were identified with pleiotropic SNPs removed (**Supplementary Table 20**). Our findings provide no support for the hypothesis that C3 protein concentration is related to the risk of the neuropsychiatric nor autoimmune disorders examined in this study.

### C3 and C4 phenome-wide association studies in the UK Biobank

With respect to C4 concentration, the PheWAS study in the UK Biobank identified significant associations with 35 phenotypes (**Supplementary Table 22 and Supplementary Figure 13**). Many of these were related to autoimmunity. Of the top 8 disease associations ranked by *P*-value, higher C4 concentration was associated with a reduced risk of six disorders, two were associated with an increased risk. The top 8 were intestinal malabsorption (which includes coeliac disease; ICD10 = K90, OR = 0.54, 95% confidence interval [CI] = 0.53 - 0.56); thyrotoxicosis [hyperthyroidism] (ICD10 = E05, OR = 0.77, 95% CI = 0.75 - 0.79); hypothyroidism (ICD10= E03, OR= 0.92, 95% CI = 0.90 – 0.93); insulin-dependent diabetes mellitus (ICD10 = E10, OR = 0.80, 95% CIs = 0.78 – 0.83); sarcoidosis (ICD10 = D86; OR = 0.79, 95% CIs = 0.75 – 0.83); psoriasis (ICD10 = L40; OR = 1.08, 95% CIs = 1.06 – 1.10); systemic lupus erythematosus (ICD10 = M32, OR = 0.74, 95% CI = 0.69 - 0.80) and ankylosing spondylitis (ICD10 = M45, OR = 1.22, 95% CIs = 1.16- 1.28). We also found a significant result for multiple sclerosis (ICD-10 = G35, OR = 0.88, 95% CI = 0.84 - 0.92). The attenuated results for the autoimmune disorders that were included in GMSR may be related to the low prevalence of these disorders (and the smaller number of cases in the UKB). In addition to the disorders which met the Bonferroni-corrected *P*-value threshold (*P*-value < 7.3×10^−6^), we note that Sjögren’s syndrome (ICD10 = M35; OR = 0.95, 95% CI = 0.92 - 0.97) was nominally significant. There were no significant findings between C4 and any neuropsychiatric disorders (ICD10 F codes). No significant difference was found between males and females. Overall, these findings lend weight to the hypotheses that neonatal C4 protein concentration is (a) not associated with the risk of neuropsychiatric disorders, but (b) is associated with reduced risks of several autoimmune disorders. There were no significant associations between C3 and any of the 1,148 phenotypes, which was in line with our GSMR findings (**Supplementary Table 23 and Supplementary Figure 14**).

## DISCUSSION

Our findings provide new insights into the genetic architecture of C3 and C4. Reassuringly, we found a robust association between C4-related haplotypes (including copy number) and neonatal C4 protein concentration. The C3 and C4 protein concentrations were phenotypically and genetically correlated (*r*_p_ = 0.65, *P*-value < 1×10^−100^; *r*_g_ = 0.35, *P*-value = 1.9×10^−35^, using all SNPs; and *r*_g_ = 0.78, *P*-value = 4×10^−5^ using trans-chr SNPs). The C3 and C4 GWAS findings identified variants in genes that encode important proteins within the inter-connected complement pathways. In contrast to a previous study^9^, we found that neither a higher imputed *C4* haplotype count nor a higher observed C4 protein concentration was associated with an increased risk of schizophrenia or any other mental disorder diagnosed in later life. We did, however, find evidence from Mendelian randomization studies that support the hypothesis that C4 protein concentration is associated with a range of autoimmune disorders. In models that incorporate the correlation between C4 and C3 protein concentrations, we found no association between C3 and an altered risk of any mental disorder nor autoimmune disorder. This following discussion will focus on 5 key findings.

First, with respect to C4 protein concentration, we found a stronger contribution of the *C4A* count compared to the *C4B* count. In keeping with prior smaller studies^18,19^, we confirmed that the copy numbers of *C4A* and *C4B* were robustly positively associated with the concentration of the C4 protein (*C4A* count; *b* = 0.33, SE = 0.005, *P*-value < 1×10^−100^: *C4B* count; *b* = 0.18, SE = 0.007, *P*-value < 1×10^−100^). In joint analyses that accounted for the pattern of correlations between the different types of *C4* allele counts, the *C4A* count had twice the contribution to overall C4 concentration compared with *C4B* count.

Second, the protein concentrations of C3 and C4, which are key components of the complement initiation pathways^82^, were highly heritable. Both pedigree- and SNP-based *h*^*2*^ estimates (standard error) were appreciable for C4 (0.40 (0.03) and 0.26 (0.006) respectively). The same estimates for C3 were smaller; 0.21 (0.03) and 0.04 (0.005) respectively. As expected, SNPs within and near the respective coding genes (*C4, C3*) contributed to more than half of the genetic variance of their related proteins.

Third, our sample sizes for C3 and C4 concentrations GWASs were nearly twenty times larger than the only published GWAS for these proteins^18^. With respect to C4 protein concentration, our series of linked GWASs provide important information about the genetic correlates of C4 protein concentration that lie outside of the MHC complex. In the GWAS, 30 quasi-independent hits were on chromosome six, within the MHC region. Six additional loci were found on chromosomes 1, 7, 9, 12, 14 and X. We identified a locus on chromosome 1 within *C4BPA*, which encodes C4 binding protein (closely involved in C4 protein regulation). The locus on chromosome 12 (rs11064501) is adjacent to two genes that encode proteins involved in complement cascade initiation (C1s, C1R). Interestingly, a locus (rs12012736) was identified on the X chromosome. This locus may be one of the factors that contributed to the small sex differences found for the C3 and C4 protein concentrations and to the known sex differences in the risk of autoimmune disorders^83^.

With respect to C3 protein concentration, apart from loci within the *C3* gene (seven quasi-independent loci within this gene on chromosome 19), we found a locus within *FCGR2B* (Fc gamma receptor IIb), which encodes a receptor for the Fc region of immunoglobulin gamma complexes (IgG). The IgG complex forms part of the machinery required for the phagocytosis of immune complexes. One locus in the MHC complex was identified, which is adjacent to the *C4A* gene. In keeping with the prior GWAS^18^, we identified two loci within *CFH*, the gene that encodes complement factor H. This protein is involved in complement regulation and has been linked to several disease phenotypes (most notably, with age-related macular degeneration)^84^. *CHF* specifically regulates C3, which slows the downstream complement activation. We also found a locus within *ABO*, which was identified as having associations with over 50 other protein concentrations^46,80^, thus variants in this gene could directly or indirectly influence generic protein metabolic pathways (e.g., upstream metabolic steps, downstream protein degradation and excretion). In summary, our study has highlighted how genetic variants within several components of the complement cascades (i.e., at the systems level) could influence the concentration of key circulation proteins such as C3 and C4. We have summarized these findings in **Figure 6**.

**Figure 6.**
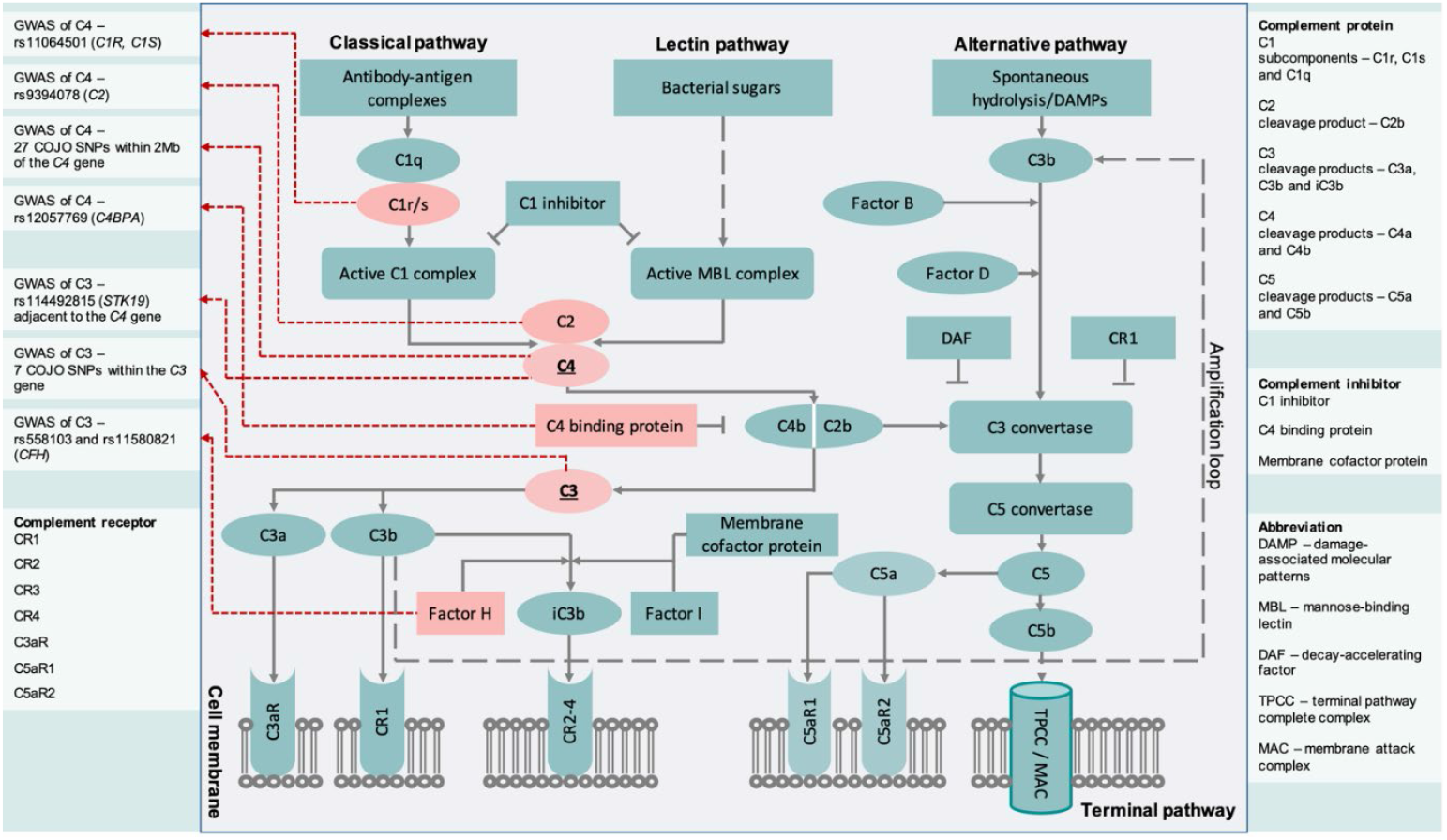
Summary of the results from GWASs of neonatal C3 and C4 protein concentrations displayed within the complement cascade. For significant loci identified from COJO, proteins encoded by annotated genes were highlighted with red colour.

Fourth, convergent evidence found no association between several *C4*-related measures and risk of SCZ. Our study measured C4 protein concentration in 68,768 neonates, an age more proximal to the period of brain development consistent with the impact of *C4* expression and synaptic pruning^6,24^. The strong association between C4-related copy number and measured C4 protein concentration, and the biologically-plausible loci/genes identified in the C3 and C4 GWASs lend weight to the validity of our measures. We found that none of the following variables were associated with an altered risk of SCZ: (a) observed neonatal C4 concentration, (b) copy numbers of either *C4A, C4B*, or *HERV*, (c) major C4-related haplotypes, nor (d) imputed brain *C4A* RNA expression. Furthermore, there were no associations between these C4-related variables and any of the other 5 iPSYCH target psychiatric disorders. We also note that the PheWAS study found no significant associations between the summary statistics of C4 protein concentration and the UK Biobank-measured brain volumes (n = 28,613). Reassuringly, we note that the SMR analyses identified (a) *C4A* gene expression was strongly linked to C4 protein concentration, and (b) higher C4 neonatal protein concentration was associated with increased *C4A* gene expression in brain tissue (including amygdala, anterior cingulate cortex, caudate basal ganglia, cerebellum, hippocampus, hypothalamus and putamen basal ganglia). These particular findings support the hypothesis that the loci we observed in the GWAS not only influence circulating C4 protein concentration (i.e., as measured in the neonatal dried blood spots), but may also influence the expression of the *C4* gene in the brain. In summary, we found no evidence to support the hypothesis that C4-related variables were causally related to the risk of SCZ, nor the other mental disorders included in the iPSYCH case-cohort study. Our findings allow us to refine future directions for schizophrenia research. We note that the most strongly associated SNPs in the two most recent SCZ GWASes (rs1233578 [28,712,247bp]^10^ and rs140365013 [27,523,869bp]^62^) are both over 3Mb upstream from the *C4A* gene (31,95 – 31.97Mb). There are 598 known genes^87,88^ (including 417 protein-coding genes) annotated between rs140365013 and the *C4A* transcription start site, so this region should provide fertile grounds for the generation of new candidate genes that might explain the top hits in recent SCZ GWASes.

Fifth, we found convergent evidence linking higher C4 protein concentration and a reduced risk of several autoimmune disorders, and an increased risk for Crohn’s disease. Based on Mendelian randomization analyses, there was robust evidence with respect to a lower risk of systemic lupus erythematous. The protective effect remained significant when we adjusted C4 concentration for MHC SNPs (from C4 with MHC SNPs to SLE: OR = 0.37, 95% CI = 0.34 – 0.42; from C4 adjusted for MHC SNPs to SLE: OR = 0.24, 95% CI = 0.12 - 0.47). Evidence also emerged for a lower risk of type-1 diabetes, multiple sclerosis, and rheumatoid arthritis. Reassuringly, the UKB-based PheWAS found variants associated with increased neonatal C4 protein concentration were associated with (a) reduced risks of a wide range of disorders (including coeliac disease, thyrotoxicosis, hypothyroidism, type 1 diabetes, sarcoidosis, SLE, nephrotic syndrome, and multiple sclerosis; Sjögren’s syndrome. was nominally significant), and (b) increased risks of several disorders (including psoriasis, ankylosing spondylitis, iridocyclitis; Crohn’s disease was nominally significant). Our findings are consistent with a meta-analysis based on 16 case-control studies, where low C4 gene copy number (<4) was associated with an increased risk of any type of autoimmune disorder, including systemic lupus erythematous^15^. A study based on large-scale genetic and transcriptomic datasets by Kim et al.^17^ suggested that *C4A-*related gene expression was not associated with risk of schizophrenia-related synaptic gene expression, but was associated with disorders including inflammatory bowel disease, rheumatoid arthritis, and lupus. Our findings support these conclusions. Recently, it was reported that variants in *C4A* and *C4B*, which were thought to increase the risk for SCZ, are protective for two autoimmune disorders (systemic lupus erythematosus, Sjögren’s syndrome)^14^. We also observed that higher C4 protein concentration was associated with increased risks of several autoimmune disorders. The mechanisms of action underpinning the pattern of increased and decreased risk of different autoimmune disorders remain poorly understood^87,88^. In summary, our findings provide convergent evidence to support the hypothesis that C4 protein concentration is associated with the risk of a range of autoimmune disorders.

Many GWASs have found links between loci in the MHC region and risk of autoimmune disorders^89-94^. Until recently, this has been interpreted as a connection between HLA genes and autoimmune disease. Recently, Kamitaki et al. have shown that the link between the MHC locus and SLE and Sjögrens may be explained by *C4A*-*C4B* allelic variance^14^ within the MHC region, thus expanding on smaller studies linking low copy number of *C4*^15,95^ and *C4A*^96^ with higher risk of systemic lupus erythematous. This raises the possibility that the correlation between the MHC region and the other above-mentioned autoimmune diseases are also explained by the *C4A*-*C4B* allelic variance. Bian et al.^41^ observed strong linkage disequilibrium with *HLA* alleles and BS (one of the *C4* alleles). Regardless of these speculations, we found no evidence of causal relationships between (a) *C4* copy number and *C4* haplotypes, (b) predicted *C4A* and *C4B* gene expressions, and (c) C4 protein concentration versus SCZ and a range of mental disorders. We identified pleiotropy between C4 concentration and three mental disorders (SCZ, DEP, BIP) from GSMR analyses. The complex linkage disequilibrium between the *C4* gene and other genes in MHC region (including HLA genes) suggests that we should be cautious when interpreting genotype to phenotype associations for loci within the MHC region.

### Strengths and Limitations of the study

Our study has several strengths. Our sample was nearly 20 times larger than the only other published GWAS of C3 and C4^18^. With respect to the hypothesis linking complement to brain development, our complement assays were collected from neonatal samples (versus adult samples). Because the onset of mental disorders such as SCZ is often in the second and third decade of life, our samples are unlikely to be impacted by reverse causation (e.g., smoking may be linked to complement gene expression in the brain^17^) and medication effects may impact on post-mortem gene expression studies^97^. With respect to limitations, because our samples were based on neonatal C3 and C4 concentrations, it remains to be seen if the genetic correlates we identified are stable across the lifespan. Also, we used an antibody that has been demonstrated to measure total C4 (i.e., both C4A and C4B), thus we are unable to isolate the concentrations of the two isoforms. The C3 and C4 concentrations in our study were derived from circulating plasma proteins, whereas the concentration of these proteins may vary between organs/tissues and also in response to local tissue activation pathways.

The study by Sekar et al.^9^, which was based on C4 haplotypes imputed from 28,799 schizophrenia cases and 35,986 controls, found that the *AL*-*AL* haplotype was associated with an odds ratio of 1.27 compared with the *BS* allele (i.e. a 27% increased odds). Our study, based on 2,517 cases and 51,799 non-cases, and using a more informative imputation training set^9^, found no significant association for this comparison (HR = 1.14, 95% CI = 0.84 - 1.54; **Supplementary Table 16**)). Because the Sekar et al. study included more schizophrenia cases in their analyses, it is feasible that our study was underpowered to confidently detect an association between C4 haplotypes and SCZ^9^. However, we had access to the concentrations of protein product of these haplotypes from a very large sample size (n = 68,768), and thus we could estimate the variance of neonatal C4 protein concentration. Based on this observed variance, our study had 80% power to confidently detect a 25% increased risk of SCZ (OR = 1.25) by 2.46 μg/L C4 protein concentration (1 standard deviation unit). Thus, our study had sufficient power to detect the effect size previously identified by Sekar et al.^9^.

## Conclusions

Our study provides new insights into the genetic and phenotypic correlates of C3 and C4 protein concentration and helps unravel the contribution of different C4-related copy numbers and haplotypes to C4 protein concentration. Based on convergent evidence, we found no evidence to support an association between C4-related measures and risk of SCZ, nor other psychiatric disorders. Mindful of the pitfalls of linking genotypes with phenotypes within the MHC region, we encourage the research community to continue to actively explore additional candidate loci for schizophrenia within this region. In contrast to our findings regarding mental disorders, convergent evidence emerged supporting an association between C4 protein concentration and risk of autoimmune disorders.

## URLs

PLINK2: https://www.cog-genomics.org/plink/2.0/

GCTA: https://yanglab.westlake.edu.cn/software/gcta/#Overview/

BayesR: https://cnsgenomics.com/software/gctb/#Overview/

BOLT-REML: https://alkesgroup.broadinstitute.org/BOLT-LMM/BOLT-LMM_manual.html

FUMA: https://fuma.ctglab.nl/

SMR: https://yanglab.westlake.edu.cn/software/smr/

GTEx version 8 (SMR format): https://yanglab.westlake.edu.cn/software/smr/#DataResource/

Human Protein Atlas: https://www.proteinatlas.org/

UCSC: https://genome.ucsc.edu/

## Data Availability

Owing to the sensitive nature of these data, individual level data can be accessed only through secure servers where download of individual level information is prohibited. Each scientific project must be approved before initiation, and approval is granted to a specific Danish research institution. International researchers may gain data access through collaboration with a Danish research institution. More information about getting access to the iPSYCH data can be obtained at https://ipsych.au.dk/about-ipsych/. The summary statistics from the GWAS for C3 and C4 and related analyses will be made available via the GWAS Catalogue https://www.ebi.ac.uk/gwas/ (Accession numbers to follow).

## Acknowledgements

The Genotype-Tissue Expression (GTEx) Project was supported by the Common Fund of the Office of the Director of the National Institutes of Health, and by NCI, NHGRI, NHLBI, NIDA, NIMH, and NINDS. The *cis*-eQTL data of GTEx version 8 was summarized into SMR format.

This research has been conducted using the data resource under the database of Genotypes and Phenotypes (dbGaP) study accession phs001992, and using the UK Biobank Resource under Application Number 12505. The authors thank GenomeDK and Aarhus University for providing computational resources and support that contributed to these research results.

We thank Dr. Joana A. Revez for her insightful comments on BayesR and R tools.

