## Supplementary Material for "The genetic and phenotypic correlates of neonatal Complement Component 3 and 4 protein concentrations with a focus on psychiatric and autoimmune disorders"

Supplementary Information

**Supplementary Tables**

**Supplementary Table 1 Descriptive summary of psychiatric disorders**

**Supplementary Table 2 C4 allele counts with combinations of *HERV***

**Supplementary Table 3 Allele frequencies of C4 common haplotypes**

**Supplementary Table 4 Distributions of C3 and C4 concentrations**

**Supplementary Table 5 Pedigree-based and SNP-based heritability of C3 and C4 concentration**

**Supplementary Table 6 Genetic correlation between C3 and C4 concentrations**

**Supplementary Table 7 Independent SNPs by COJO from GWAS of C4 protein concentration**

**Supplementary Table 8 Independent SNPs by COJO from GWAS of C3 protein concentration**

**Supplementary Table 9 Effects of C4 copy numbers on C3 and C4 concentrations by GREML**

**Supplementary Table 10 Joint effects of C4 haplotypes on C3 and C4 concentrations by GREML**

**Supplementary Table 11 Significant genes of C4 concentration by FUMA**

**Supplementary Table 12 SMR results of the C4 protein concentration**

**Supplementary Table 13 Significant genes of C3 concentration by FUMA**

**Supplementary Table 14 SMR results of the C3 protein concentration**

**Supplementary Table 15 Hazard ratios of C4 copy numbers on mental disorders**

**Supplementary Table 16 Hazard ratios of C4 haplotypes on mental disorders**

**Supplementary Table 17 Hazard ratios of predicted C4 gene expressions on mental disorders**

**Supplementary Table 18 Hazard ratios of protein concentrations on mental disorders**

**Supplementary Table 19 GWAS summary statistics for GSMR analyses**

**Supplementary Table 20 GSMR analyses to identify causal relationships of C3 and C4 protein concentrations**

**Supplementary Table 21 GSMR analyses to examine reverse causations for C3 and C4 protein concentrations**

**Supplementary Table 22 PheWAS analysis of the C4 protein concentration in UKB**

**Supplementary Table 23 PheWAS analysis of the C3 protein concentration in UKB**

### **SUPPLEMENTARY METHODS**

#### **Supplementary Method 1** Sample preparation steps and pre-analytical variation

These methods have been described in a related study^1^. Two 3.2 mm disks from neonatal dried blood spot (DBS) samples were punched into each well of polymerase chain reaction plates (72.1981.202, Sarstedt). 130 µL extraction buffer (PBS containing 1% BSA (Sigma Aldrich #A4503), 0.5% Tween-20 (#8.22184.0500, Merck Millipore), and complete protease inhibitor cocktail (#11836145001, Roche Diagnostics)) was added to each well, and the samples were incubated for 1 hour at room temperature on a microwell shaker set at 900 rpm. After separating the extract from the filter paper into sterile Matrix 2D tubes (#3232, ThermoFisher Scientific), the extracts were stored at -80°C for 6-7 years before analysis.

To validate the stability of the samples during storage, we randomly selected 15-16 samples stored from 5 years (1984, 1992, 2000, 2008, and 2016; in total 76 samples). After extracting the samples and adding them to an MSD plate, the rest of the extracts were frozen for 2 months, thawed and measured as described above to imitate the freeze-thaw cycle of the samples in the study. This showed (**Supplementary Figure 3**) the C3 and C4 samples from 1984 had lower levels. A likely explanation of the variation would be the change of filter paper in around 1989, where “Schleicher & Schuell grade 2992” was gradually replaced by “Schleicher & Schuell grade 903”.

#### **Supplementary Method 2** Genetic ancestry inference

There were 80,873 individuals of multiple ancestries in the iPSYCH2012 study. We projected the individuals to the 1000 Genome phase 3 v5 (1KG ph3v5) reference data^2^. The projection only included the quality controlled (QCed) SNPs from both iPSYCH2012 and 1KG reference datasets. The first two principal components (PCs) were estimated by FastPCA^3^. For the PC estimations, we further excluded the SNPs in the reported regions of long-range linkage disequilibrium (LD)^4,5^, and performed a LD pruning for the remaining SNPs, pairwise LD *r*^2^ < 0.2 within a window of 500Kb. We then conducted a k-means clustering method to partition the individuals. Participants were considered as Europeans if the posterior probabilities were > 0.9 of the European cluster in the 1KG reference data. The remaining participants were considered as non-Europeans. 75,764 individuals were assigned to European ancestry (**Supplementary Figure 1**). We only included these European individuals in the study. The genetic relationship matrix (GRM) of the European individuals was estimated by GCTA v1.93.2^6^ using the 5,201,724 common autosomal SNPs. There were 56,651 individuals with pairwise genetic relatedness < 0.05. We estimated the first 20 PCs by FastPCA for the iPSYCH2012 individuals of European ancestry using the same approach as above. In addition, we excluded SNPs in the MHC region (chr6: 24.8Mb – 33.9Mb, hg19) in addition to SNPs with long-range LD, in order to improve the power in the association studies.

#### **Supplementary Method 3** Determination of *C4* polymorphism combinations

The imputation results provide the counts of 3 *C4* alleles, *C4A*, *C4B* and *HERV*. It is unable to confidently distinguish between all combinations of variants, for example, between the haplotypes *AS-BL* and *AL-BS*. Therefore, we examined the counts of *C4* allele combinations by restricting the imputed results to those without ambiguity, which met at least one of the criteria, a) number of *C4A* allele = 0, b) number of *C4B* allele = 0, 3) no HERV and c) number of *C4* gene copy (including both *C4A* and *C4B*) is equal to number of *HERV*. The counts of all combinations were shown in **Supplementary Table 2**.

#### **Supplementary Method 4** Examining ascertainment bias in the GWASs of C3 and C4 protein concentrations

Because the iPSYCH2012 case-cohort study is enriched with individuals with mental disorders, we explored if this design feature could bias the GWAS findings for the neonatal C3 and C4 protein concentrations. First, we undertook modelling based on a set of assumptions related to potential bias. The simulation methods in detail can be found below. The simulations found that the presence of an association between the mental disorders and protein concentration could potentially introduce biases (**Supplementary Figure 8**); however, this bias does not exist if the focal trait (in this case, mental disorders) was independent of the secondary trait (in this case, C3 and C4 protein concentrations) (**Supplementary Figure 9**). To investigate this further, we conducted the GWAS analyses using two subsets of participants in the iPSYCH2012, 1) randomly sampled population-based sub-cohort (25,078 participants of European ancestry), 2) cases of six mental disorders outside the randomly sampled sub-cohort (50,686 participants of European ancestry). The GWAS estimates in the sub-cohort were consistent with those in the entire population (**Supplementary Figure 10**), and therefore the following post-GWAS analyses were based on the results from the full case-cohort sample.

*Simulation methods of ascertainment bias*

We simulated 5M individuals as an entire population. In the entire population, I simulated two scenarios 1) the secondary trait was independent of the focal trait; 2) the secondary trait was correlated with focal trait. In the first scenario, I simulated 300 common SNPs (*z*), including 100 causal variants for mental disorder (focal trait, *y*), 1-100; 100 causal variants for the secondary trait (*x*), 101-200, and 100 null SNPs, 201-300. In the second scenario, a subset of causal variants of *x* was shared with *y*, 100 causal variants for *y* (1-100), 100 causal variants for *x* (1-50, 101-150) and 150 null SNPs (151-300). Genotypes of each SNP (e.g., the *i*-th SNP) were from a binomial distribution, *z*_i_~*B*(2, *f*_i_) with *f*i being its allele frequency, *f*_i_ ≥ 0.01. The two traits, *x* and *y*, were simulated using the model based on respective causal variants, *t_j_* = Σ(*z_i_b_i_*) + e*_j_*, where *t* represents the trait of *x* or *y*; *z_i_* represents the *i*-th causal variants; *b_i_* represents the effect of the *i*-th causal variants; We simulated the effects at all 300 causal variants from a standard normal distribution *N*(0,1); *e* represents residuals, *e*~*N*(0, var(Σ(*z_i_b_i_*))×(1/*R*^2^ - 1); *R*^2^ represents the total genetic variance in trait *x* or *y* explained by respective causal variants, *R*^2^ = (for *x*) 60% and (for *y*) 40%, respectively. The simulated *x* and *y* were standardised with mean 0 and variance 1. We set the population prevalence of *y* 2%, which leaded to 100K cases and 4,900K controls. To mimic the iPSYCH2012 cohort, we constructed a case-control cohort including two sub-cohorts sampled from the entire population, 1) popu-subset1, 30K randomly sampled individuals and 2) popu-subset2, 26K randomly sampled cases. The individuals in the popu-subset1 could be overlapping with those in the popu-subset2. We then performed association studies in both the entire population and the case-control cohort. The simulation was replicated 1,000 times.

### **SUPPLEMENTARY FIGURES**

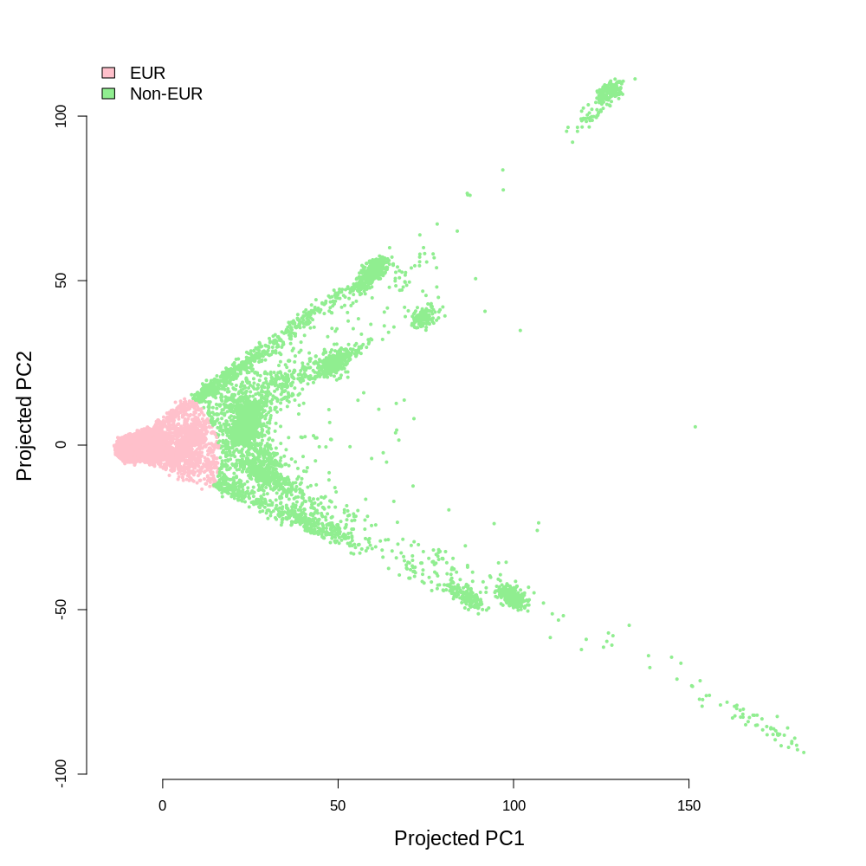

#### **Supplementary Figure 1** Ancestry inference for the iPSYCH2012 study.

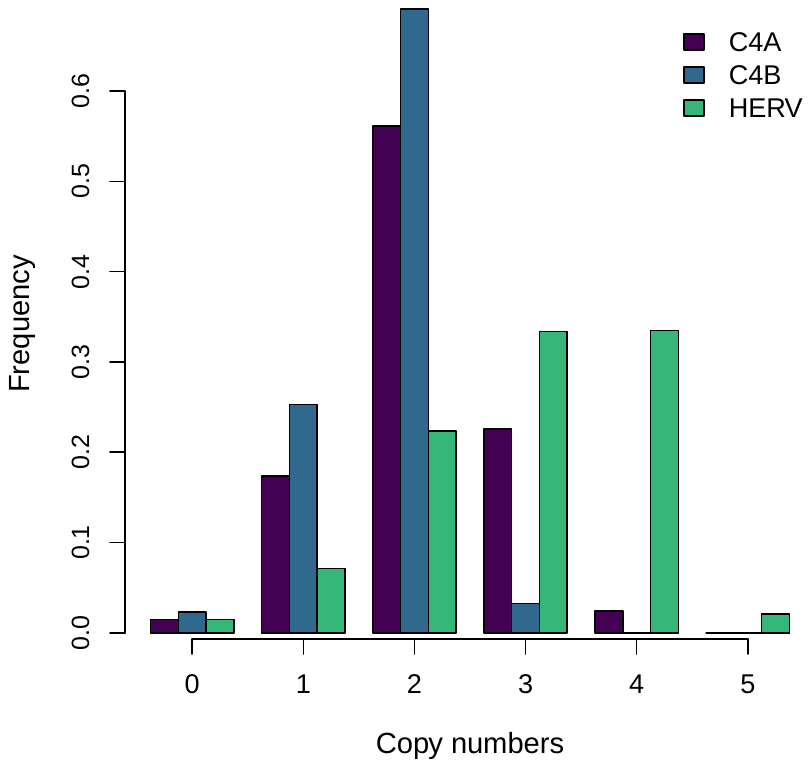

**Supplementary Figure 2** Copy numbers of C4 alleles. There were three types of C4 alleles, C4A, C4B and HERV, presented with three colours, respectively.

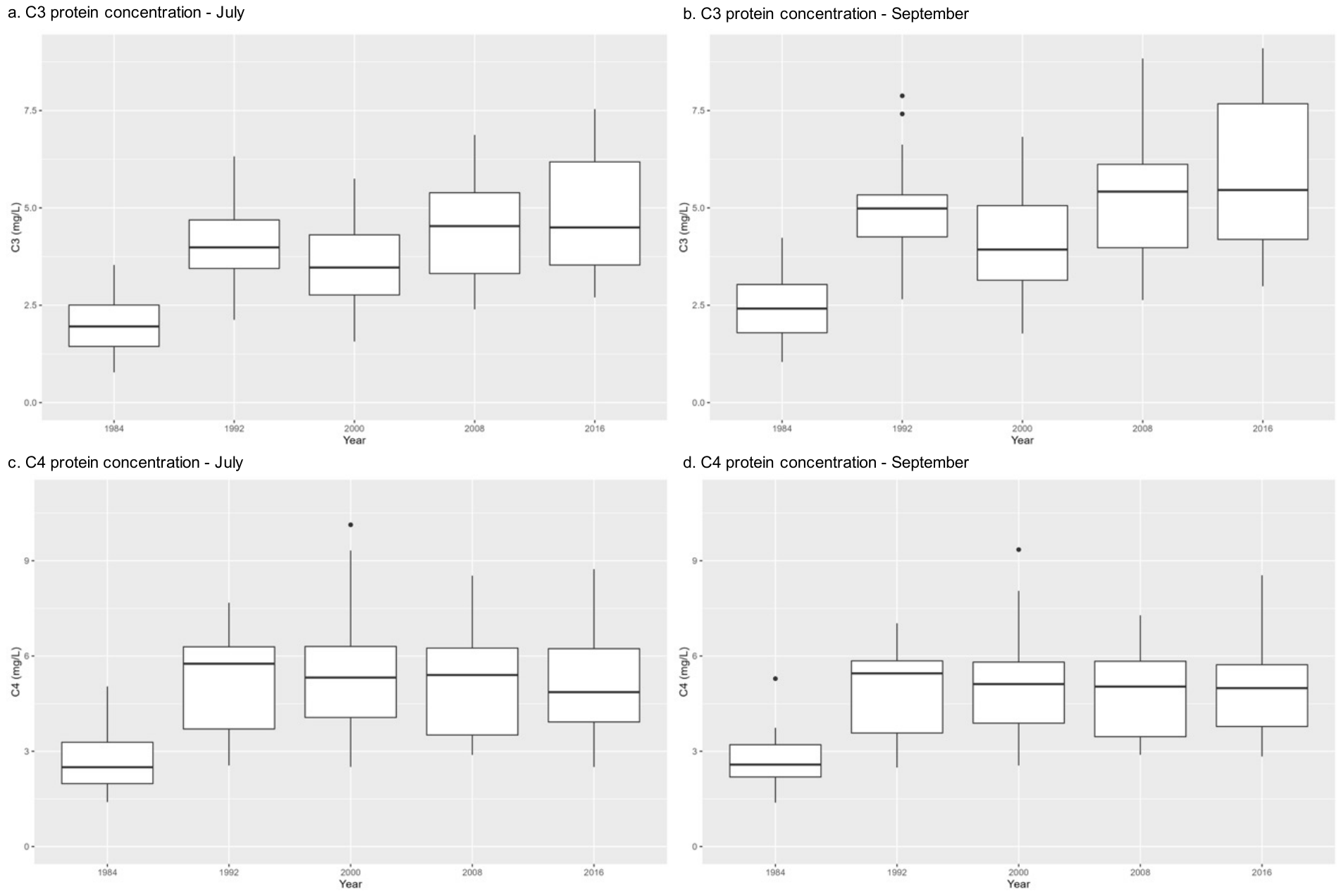

**Supplementary Figure 3** Degradation of the C3 and C4 protein concentrations. Boxplots showing the median value of the C3 (a and b) and C4 (c and d) protein concentrations split by year of birth (x-axis). The samples were extracted in July and analysed in July (a and c) and September (b and c) after 2 months storage at -80 °C. Y-axis displays concentration in mg/L. Hinges of boxes are first and third quartiles.

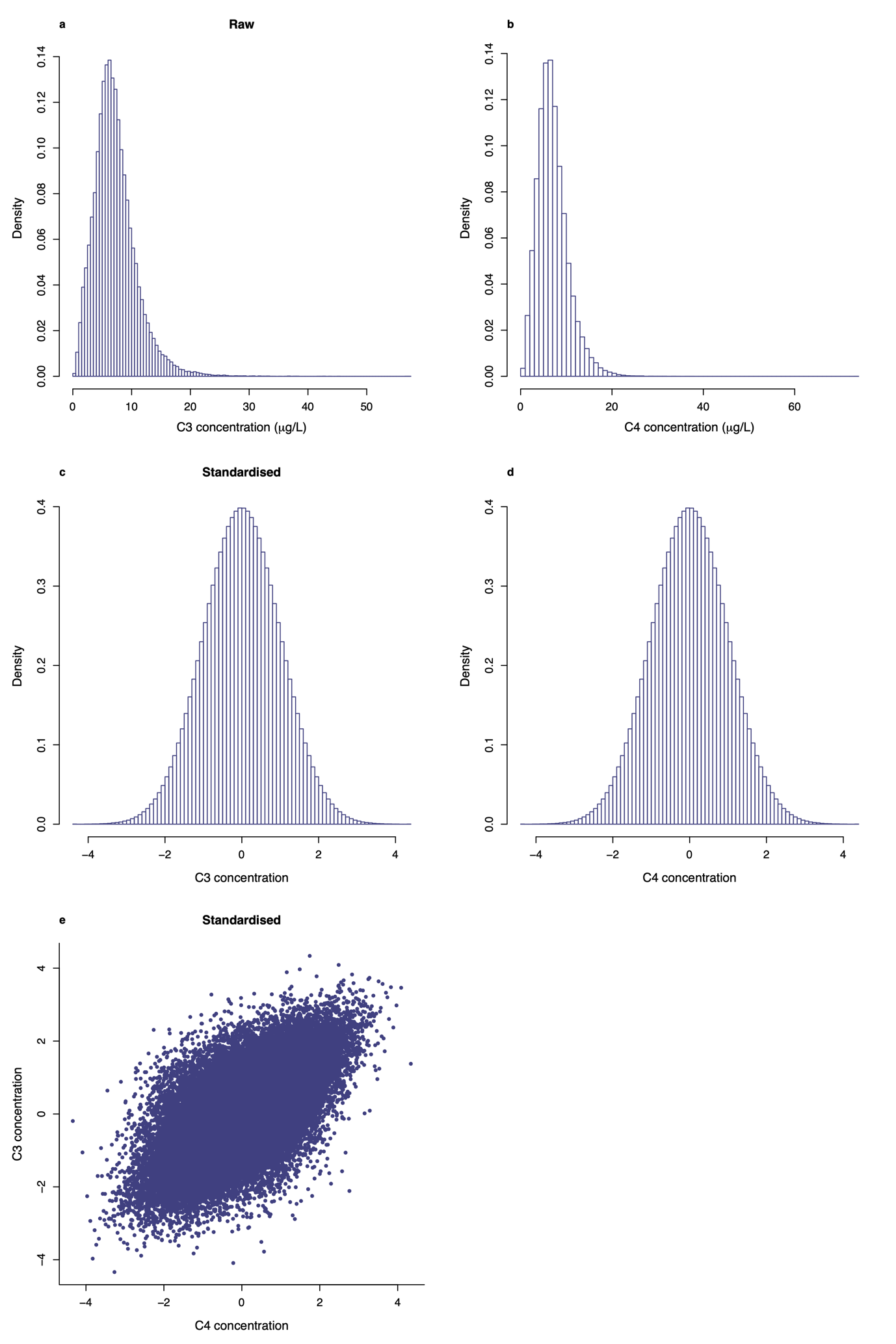

**Supplementary Figure 4** Distributions of the C3 and C4 protein concentrations. Panels in the first row represent the distributions before standardisation, (a) C3 and (b) C4 protein concentrations. The distributions after standardisation were shown in panels (c) for C3 concentration and (d) for C4 concentration. Panel (e) represents the phenotypic correlation between the two concentrations after standardisation.

a
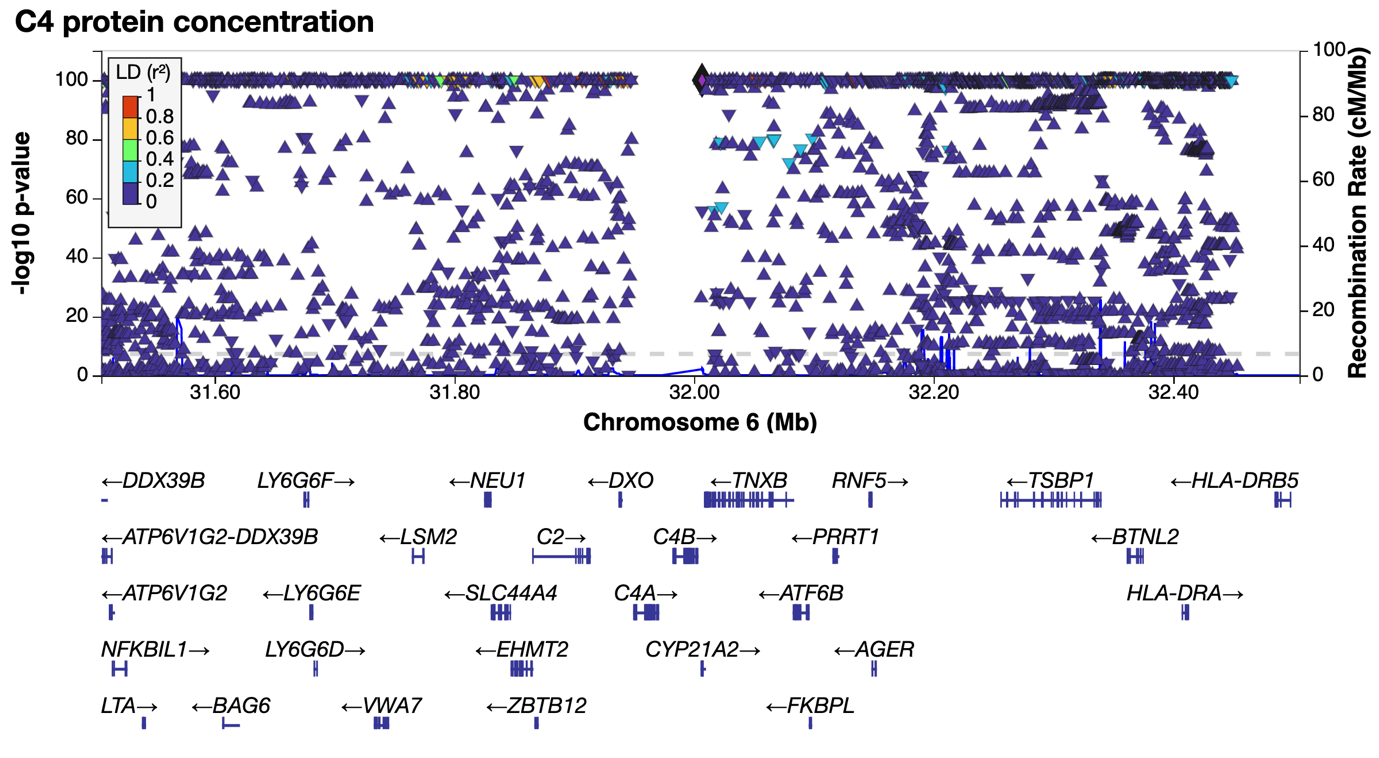

b
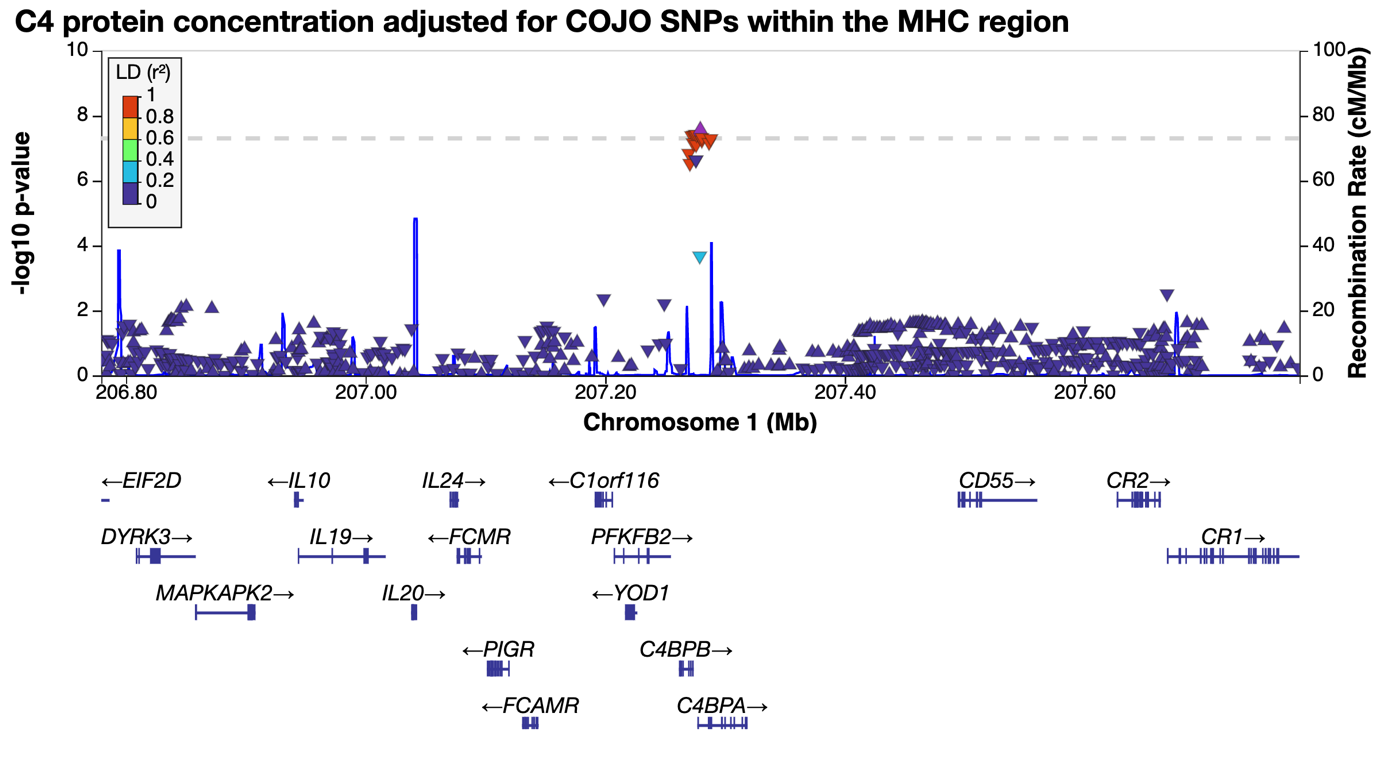

c
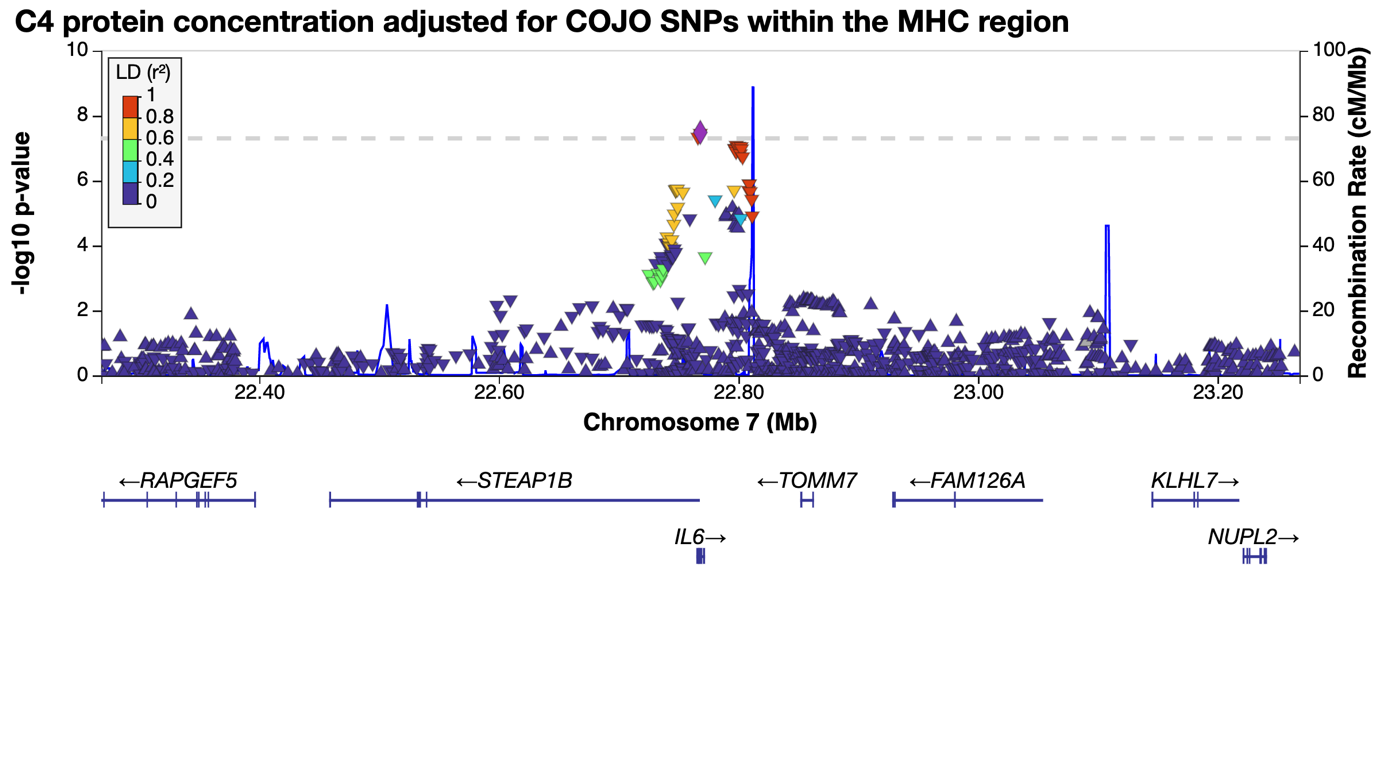

d
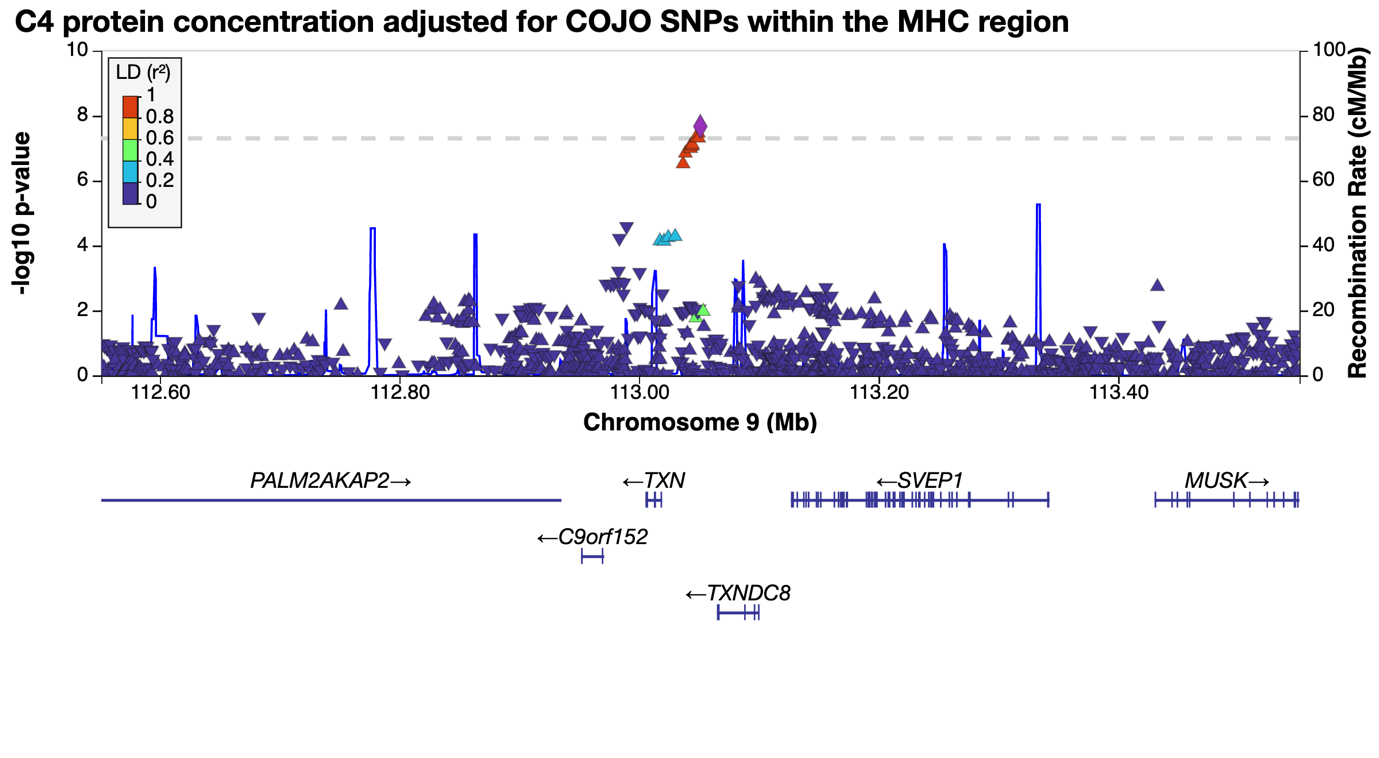

e
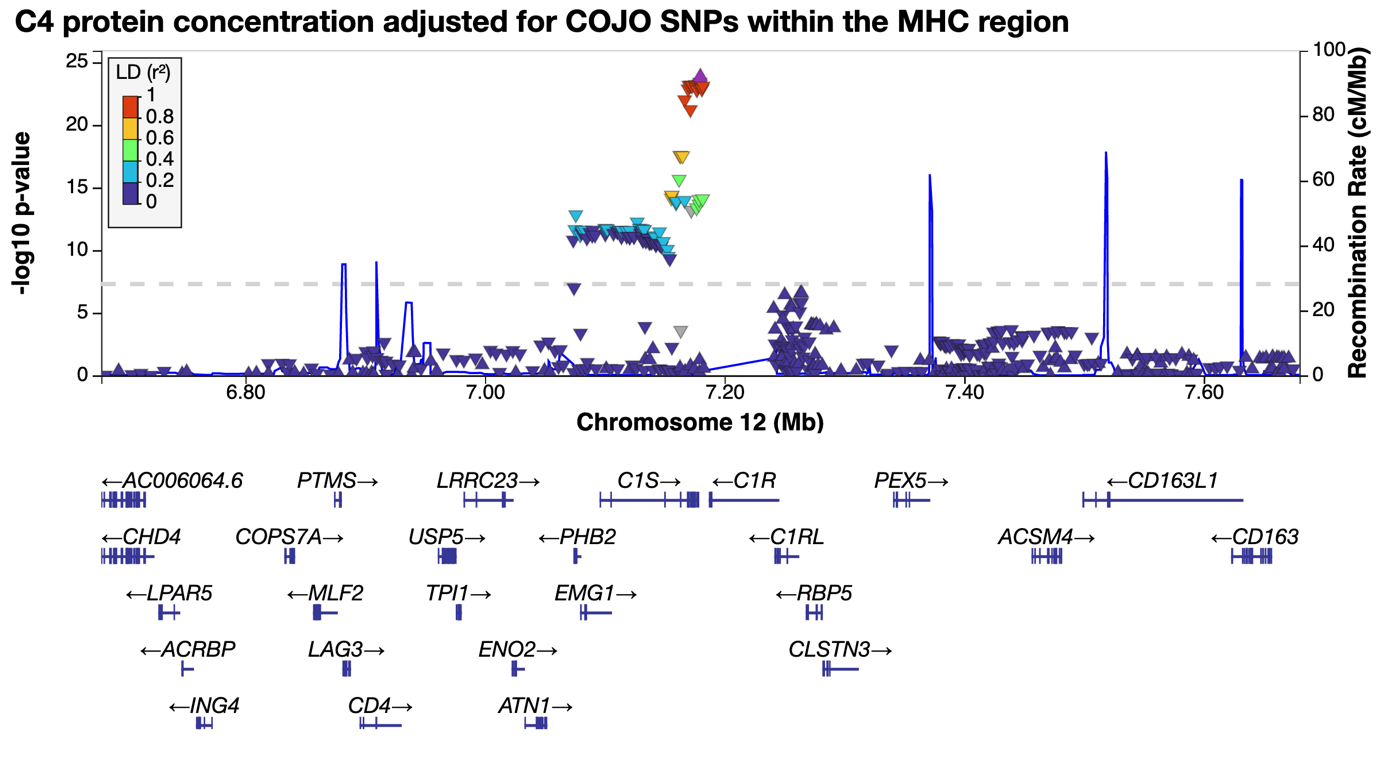

f
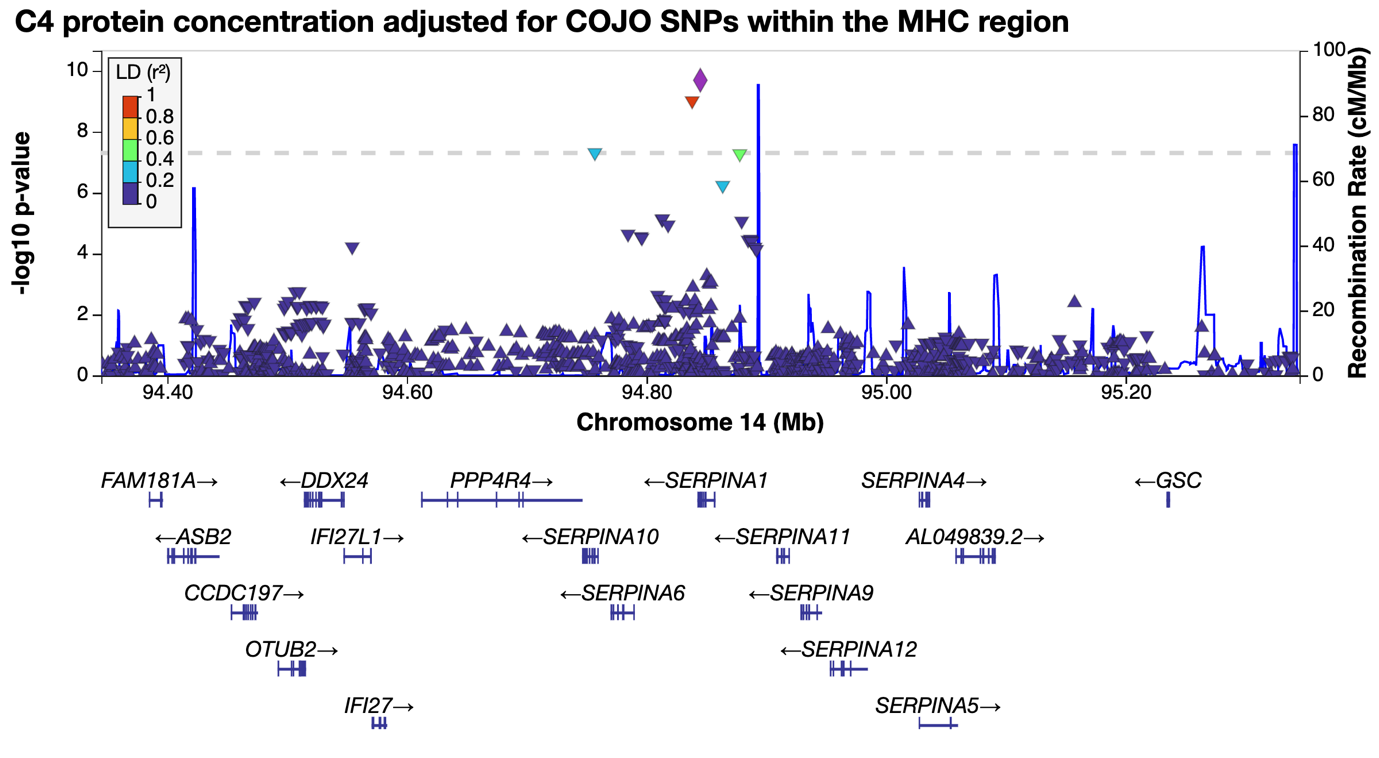

g
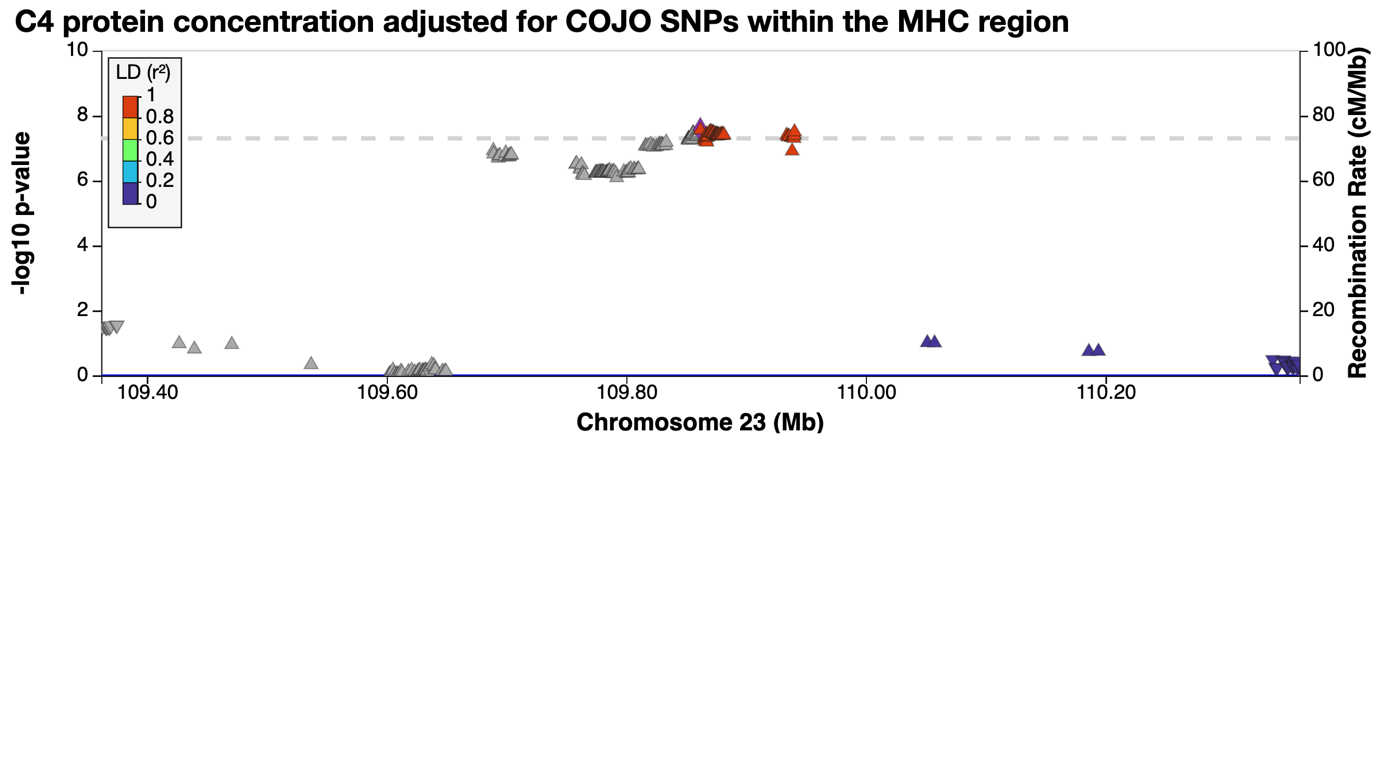

**Supplementary Figure 5** COJO loci identified from GWAS of C4 protein concentration. The top panel (panel a) showed GWAS *P*-values of SNPs in and near the *C4A* and *C4B* genes on chromosome 6—for this panel the -log P-value shown on the vertical axis has been truncated at 50 for graphical presentation. The following panels show the 6 COJO loci on 6 chromosomes, chromosome 1 (panel b), chromosome 7 (panel c), chromosome 9 (panel d), chromosome 12 (panel e), chromosome 14 (panel f) and chromosome X (panel g). The *P*-values shown in the top panel were from GWAS of unadjusted C4 protein concentrations. The *P*-values of the remaining 5 loci were from GWAS of C4 protein concentration adjusted for COJO SNPs in and near MHC region.

a
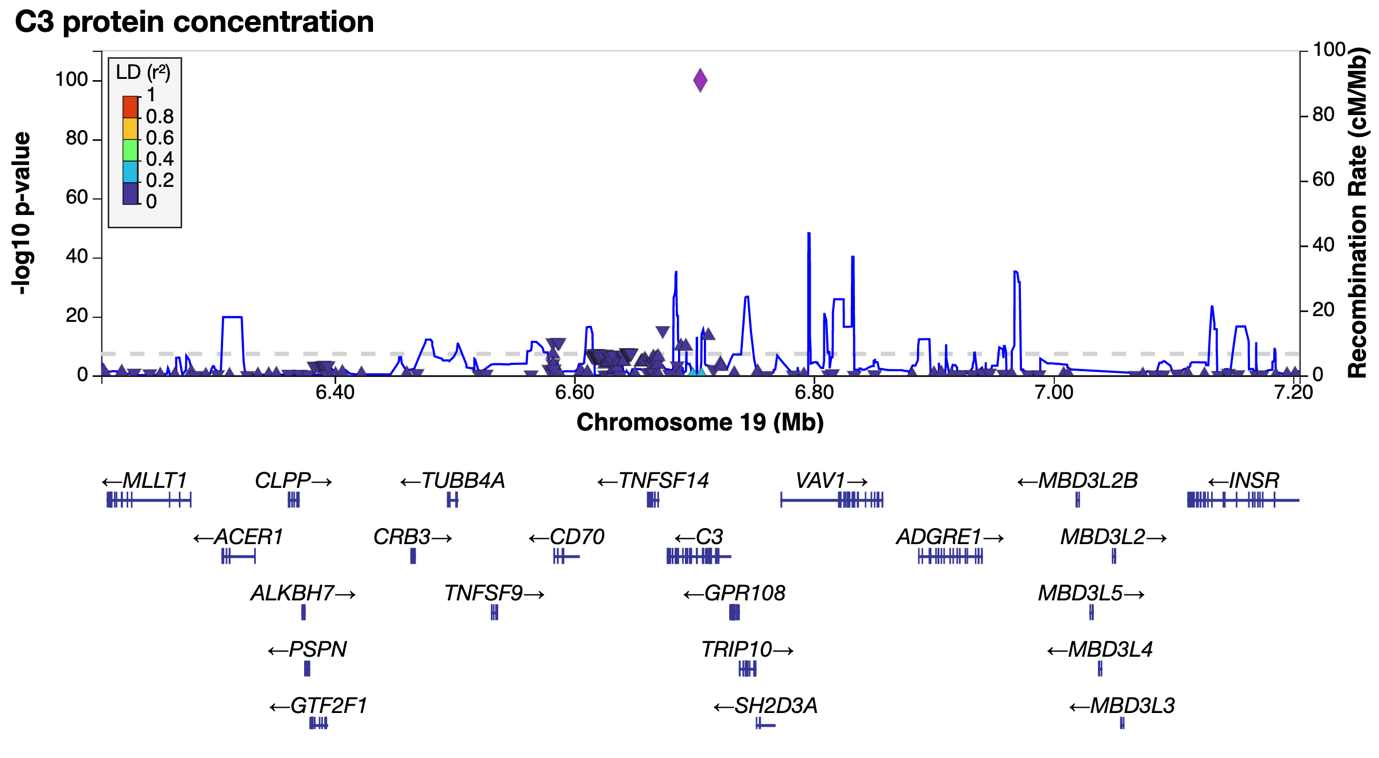

b
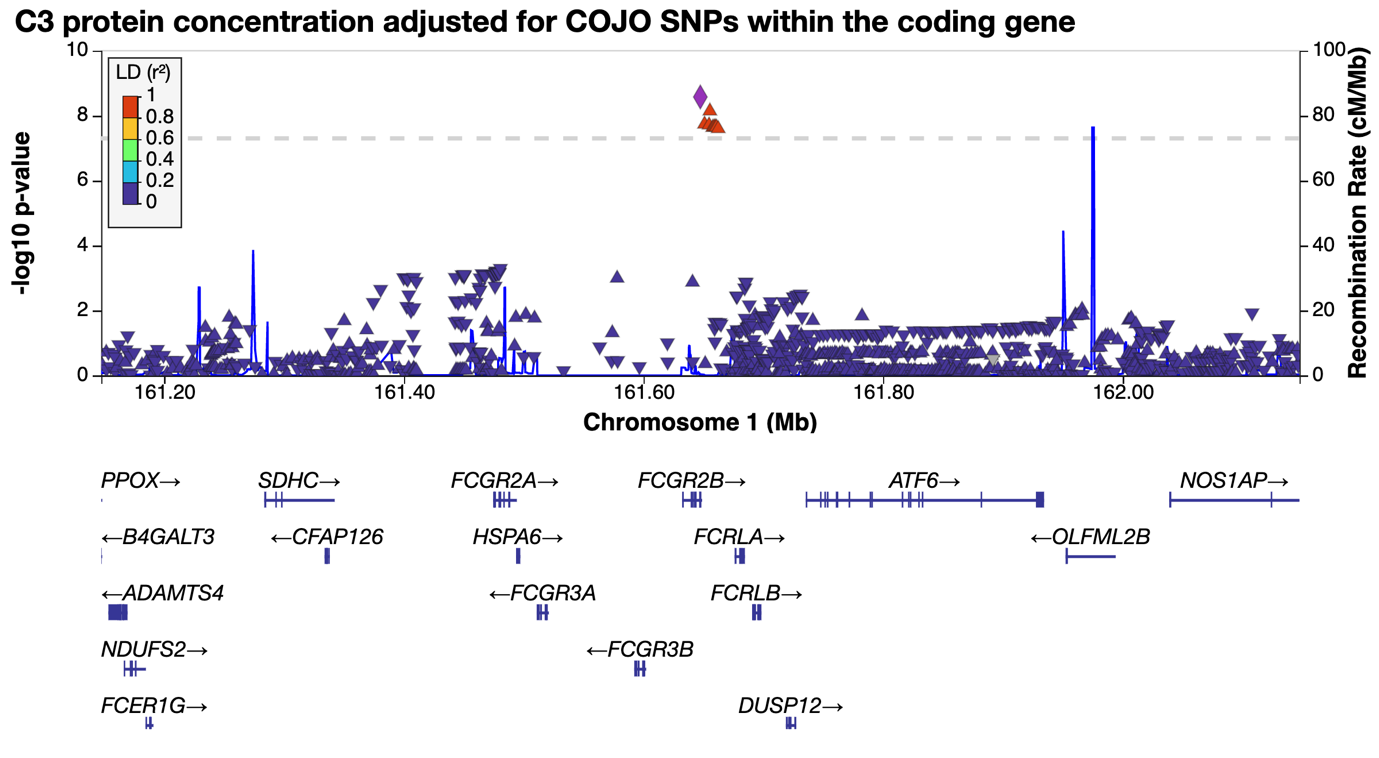

c
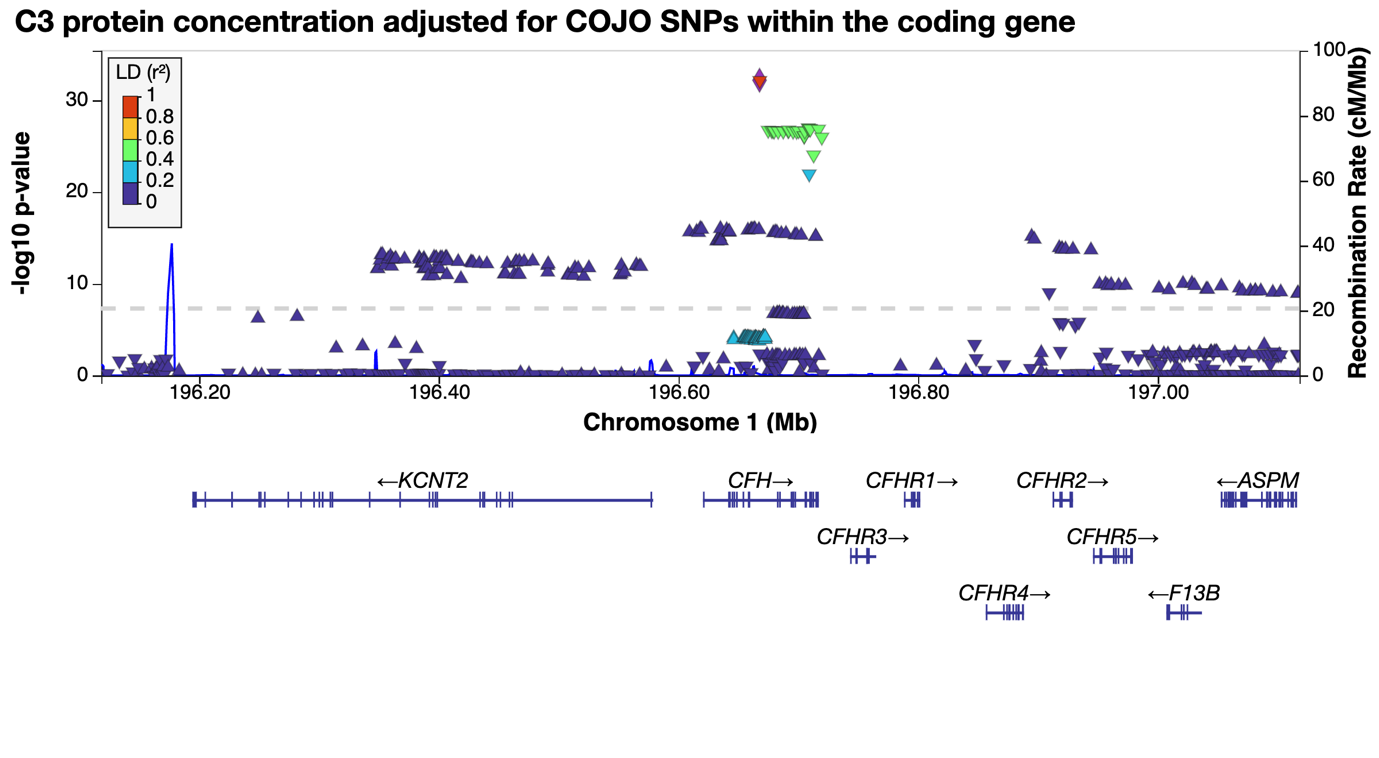

d
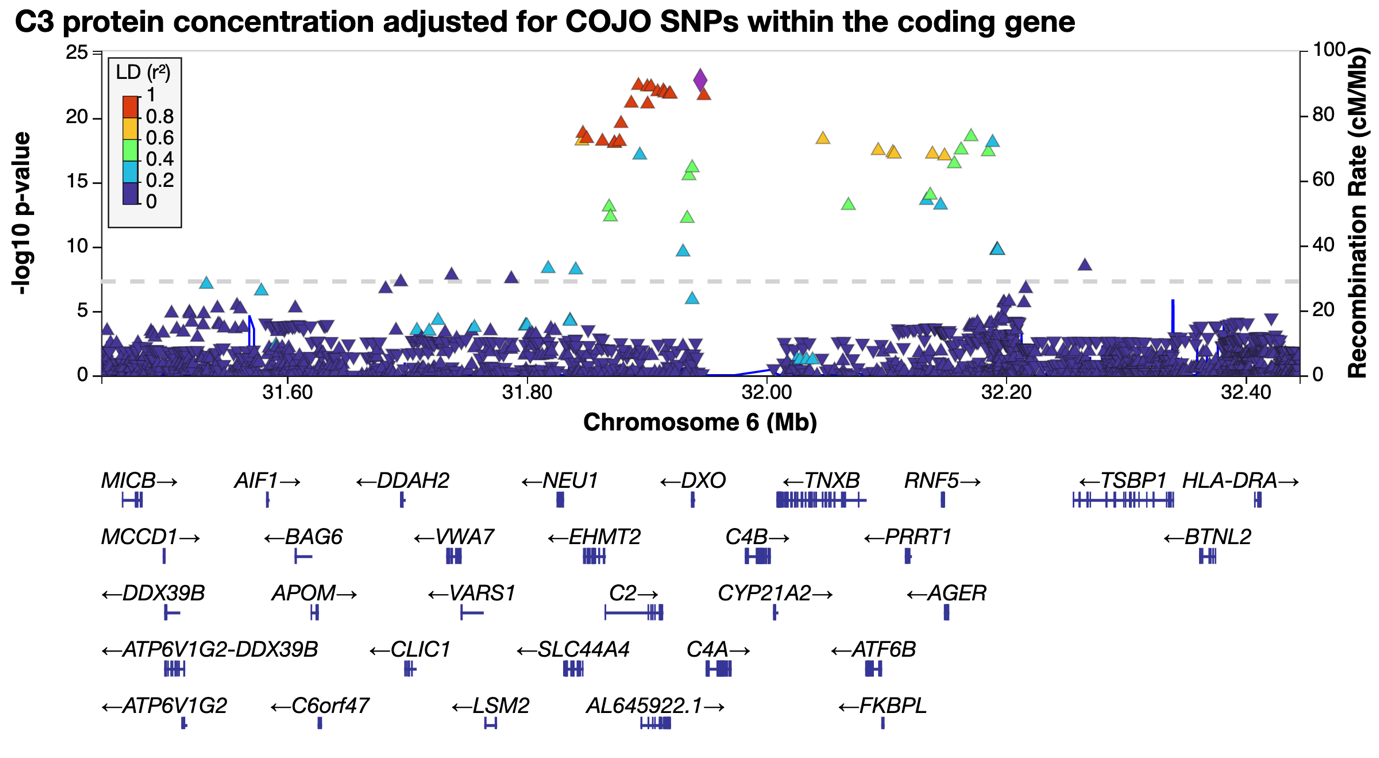

e
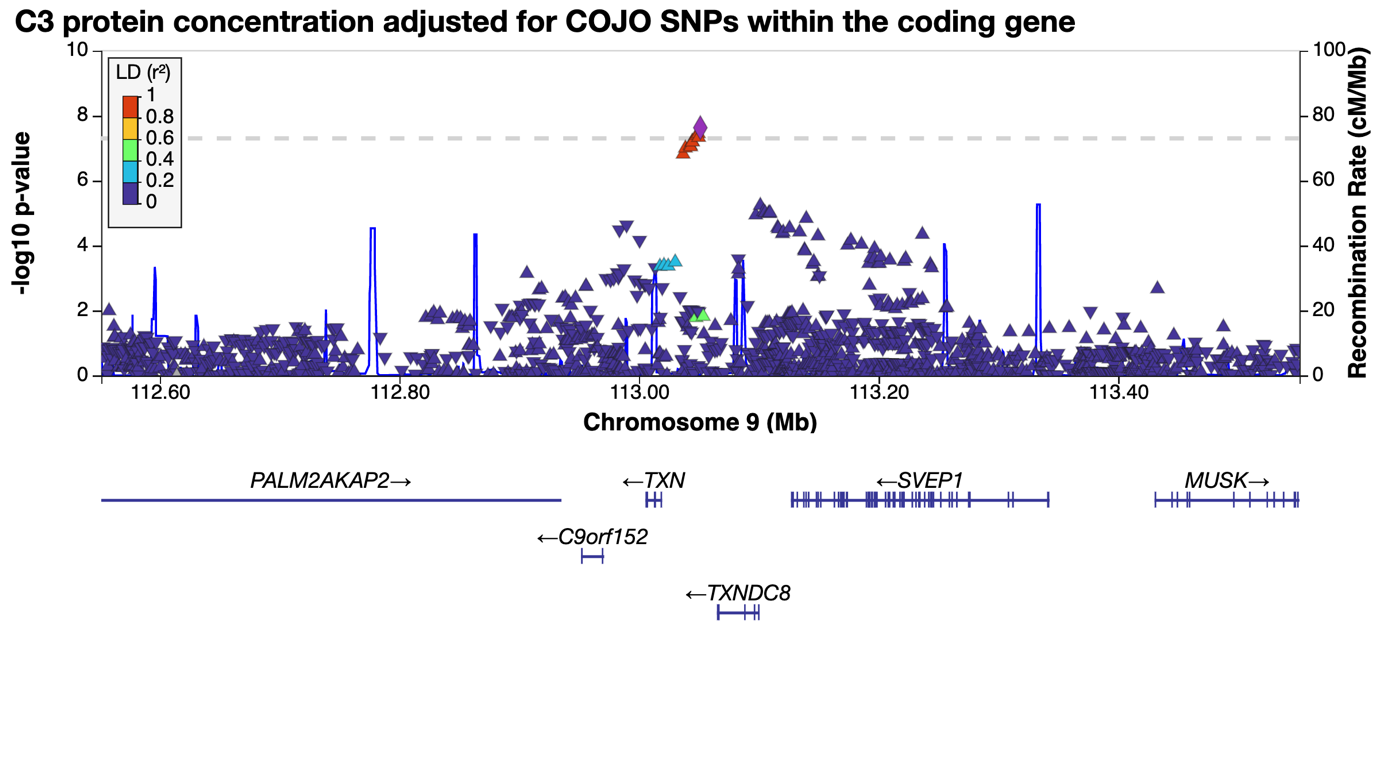

f
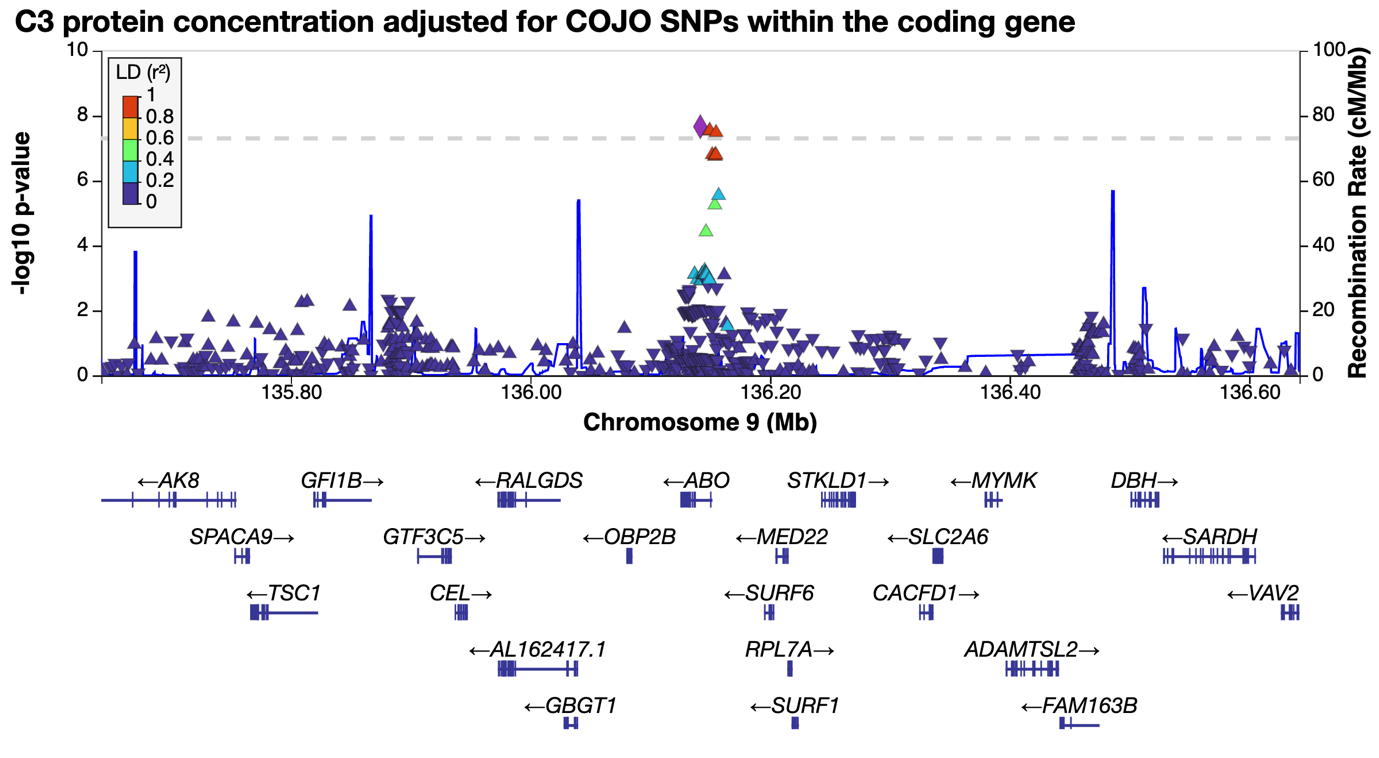

g
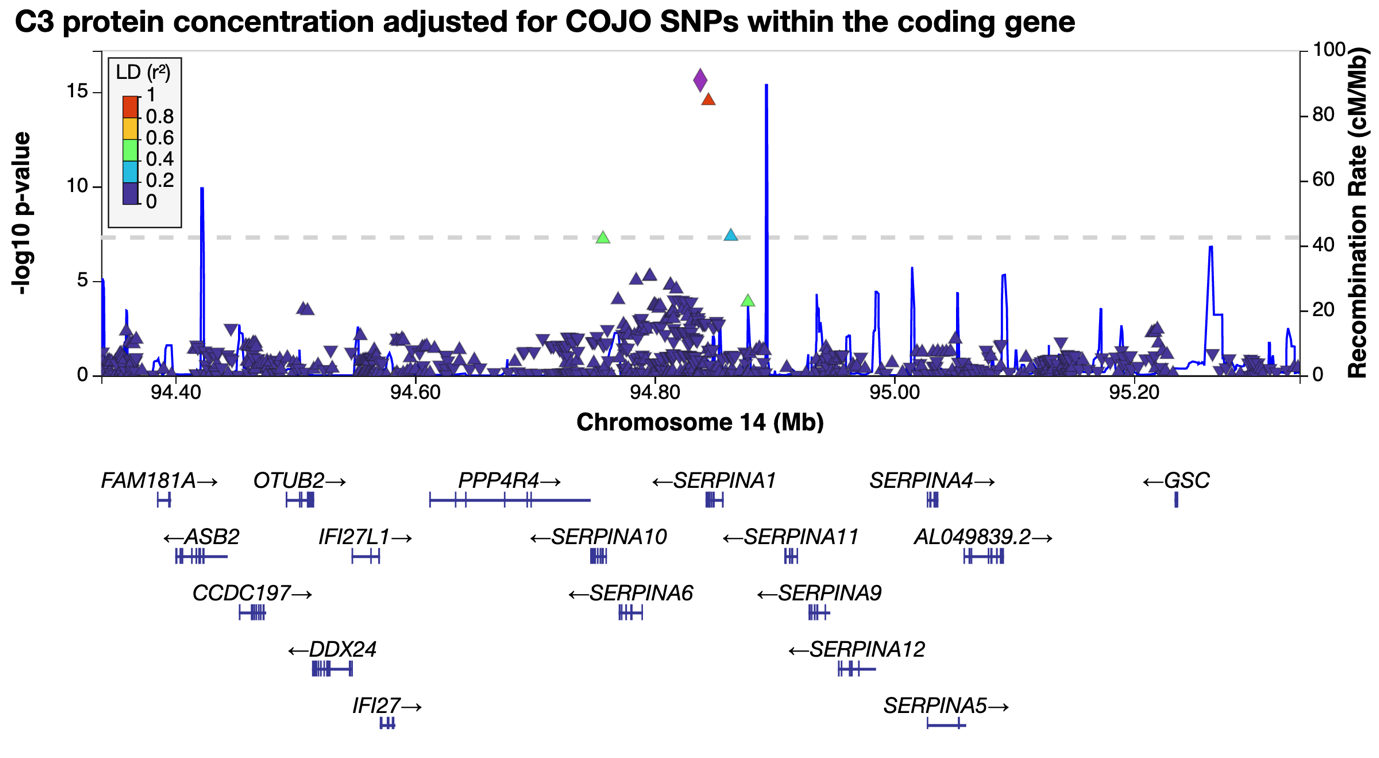

h
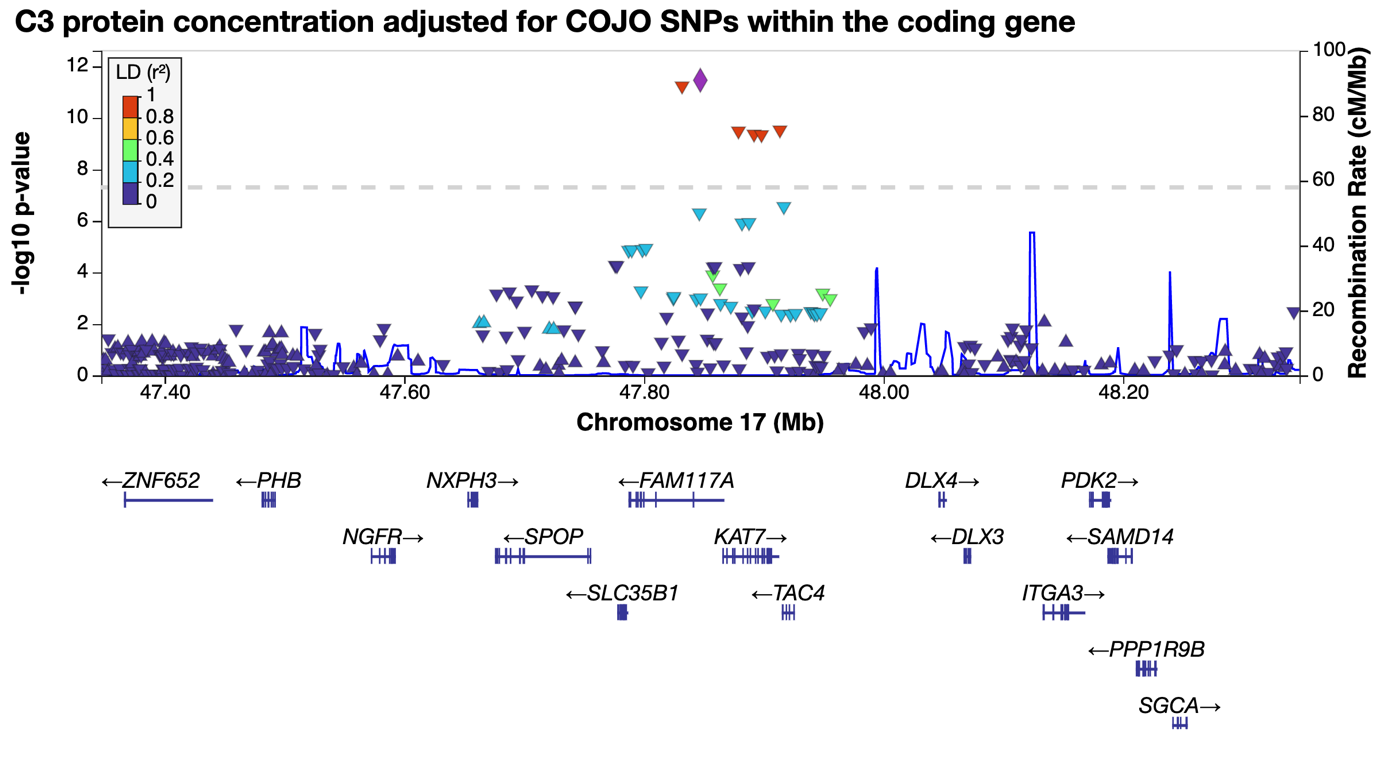

**Supplementary Figure 6** COJO loci identified from GWAS of C3 protein concentration. The top panel (panel a) showed GWAS *P*-values of SNPs in and near the *C3* gene on chromosome 19. The following panels showed the 8 COJO loci on the 5 chromosomes, chromosome 1 (panels b-c), chromosome 6 (panel d), chromosome 9 (panels e and f), chromosome 14 (panel g) and chromosome 17 (panel h). The *P*-values in the top panel were from GWAS of unadjusted C3 protein concentration. The *P*-values of the remaining 8 loci (panels b-h) were from GWAS of C3 protein concentrations adjusted for COJO SNPs on chr19.

a) C4 concentration

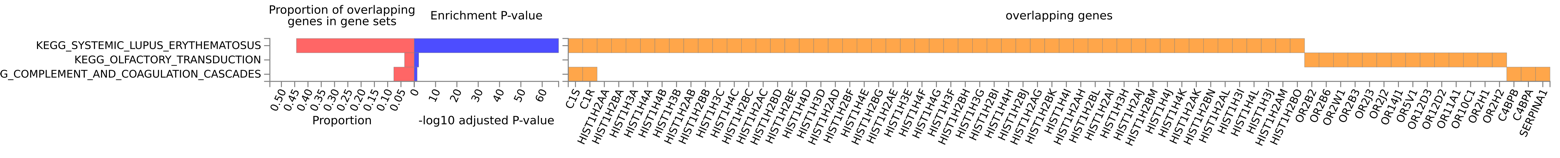

b) C3 concentration

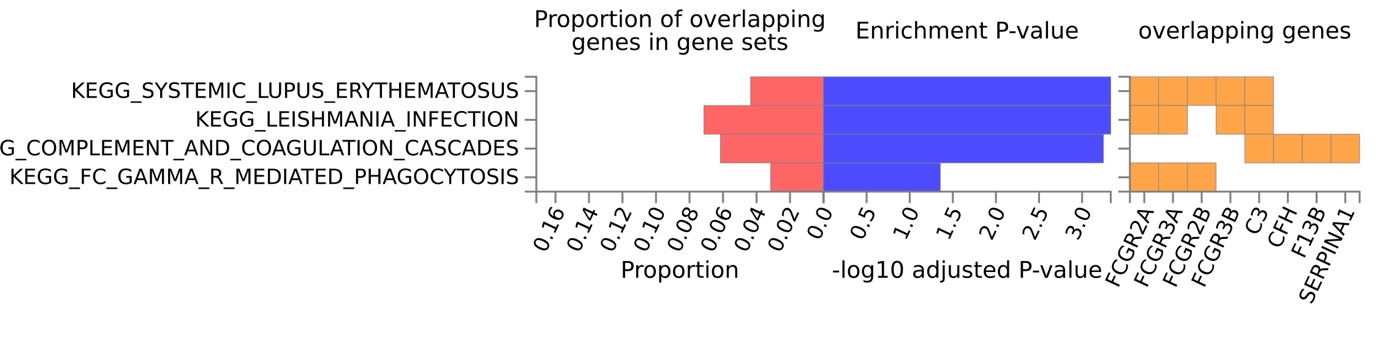

#### **Supplementary Figure 7** Enrichment analysis of protein concentrations with KEGG gene-sets

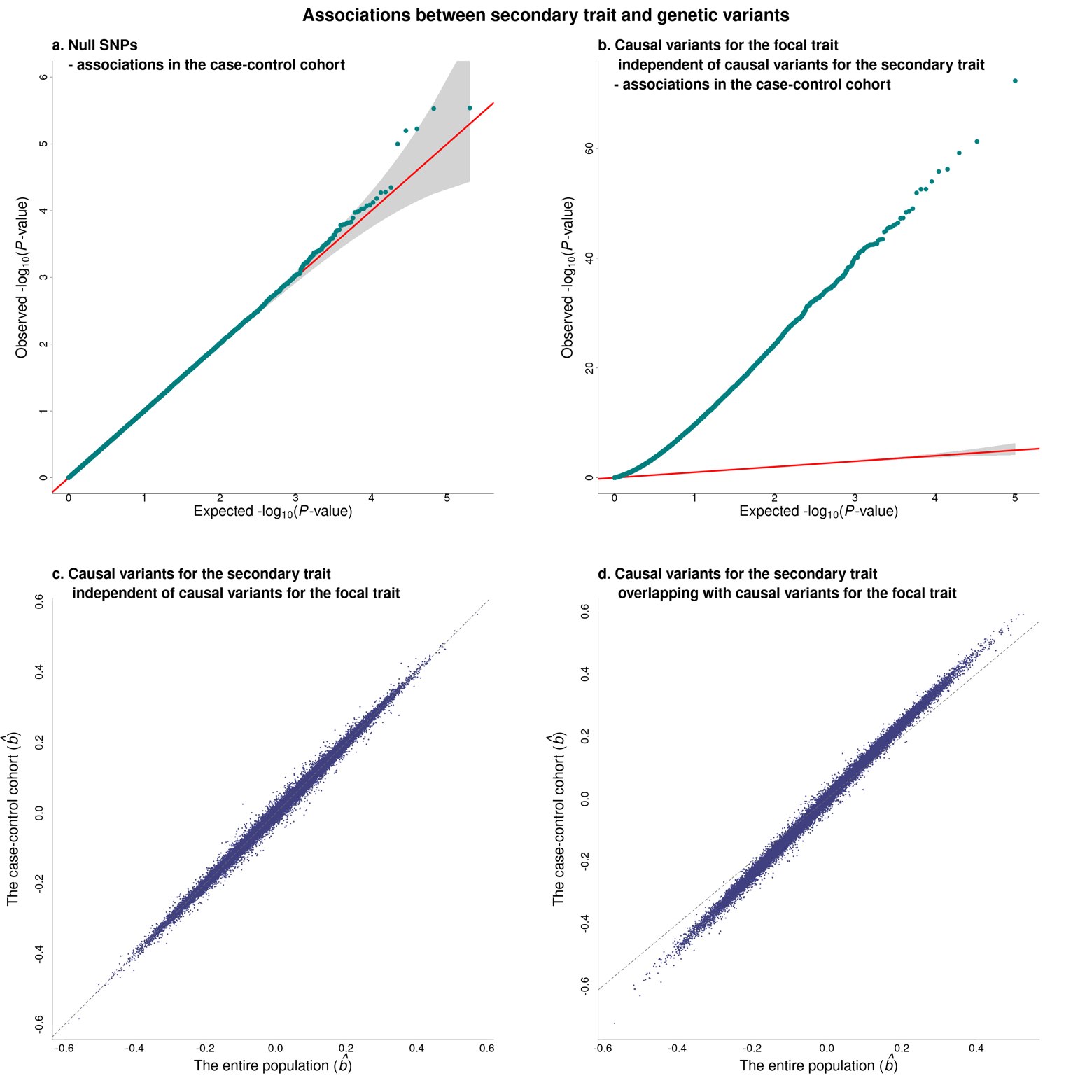

**Supplementary Figure 8** Associations between secondary trait and genetic variants when the secondary trait is associated with the focal trait. The simulation details can be found in Supplementary Methods #3. In brief, we randomly sampled a cohort with 5M individuals as an entire population. In the population, we simulated 300 variants in total, including 100 causal variants for mental disorder (the focal trait, *y*), 100 causal variants for the secondary trait (*x*) and 150 null SNPs. 50 causal variants were shared by both *x* and *y*. Assuming the disease prevalence of *y* was 2%, there were 100K cases and 4,900K non-cases. To mimic the iPSYCH2012 cohort, a case-control cohort was sampled from the entire population with two sub-cohorts 1) 30K randomly sampled individuals and 2) 26K randomly sampled cases. Duplicated samples were removed. Association analyses between the secondary trait and 300 causal variants were conducted in both the entire population and the case-control cohorts. Shown in the plot were estimates of the three sets of SNPs in the association analyses, (a) QQ plot of the 150 null SNPs, *P*-values from associations in the case-control cohort, (b) QQ plot of the 50 causal variants *only* associated with *y*, *P*-values from associations in the case-control cohort, (c) for the 50 causal variants *only* associated with *x*, comparison of effects between the entire population and the case-control cohort, and (d) for the 50 causal variants shared by *x* and *y*, comparison of effects between the entire population and the case-control cohort.

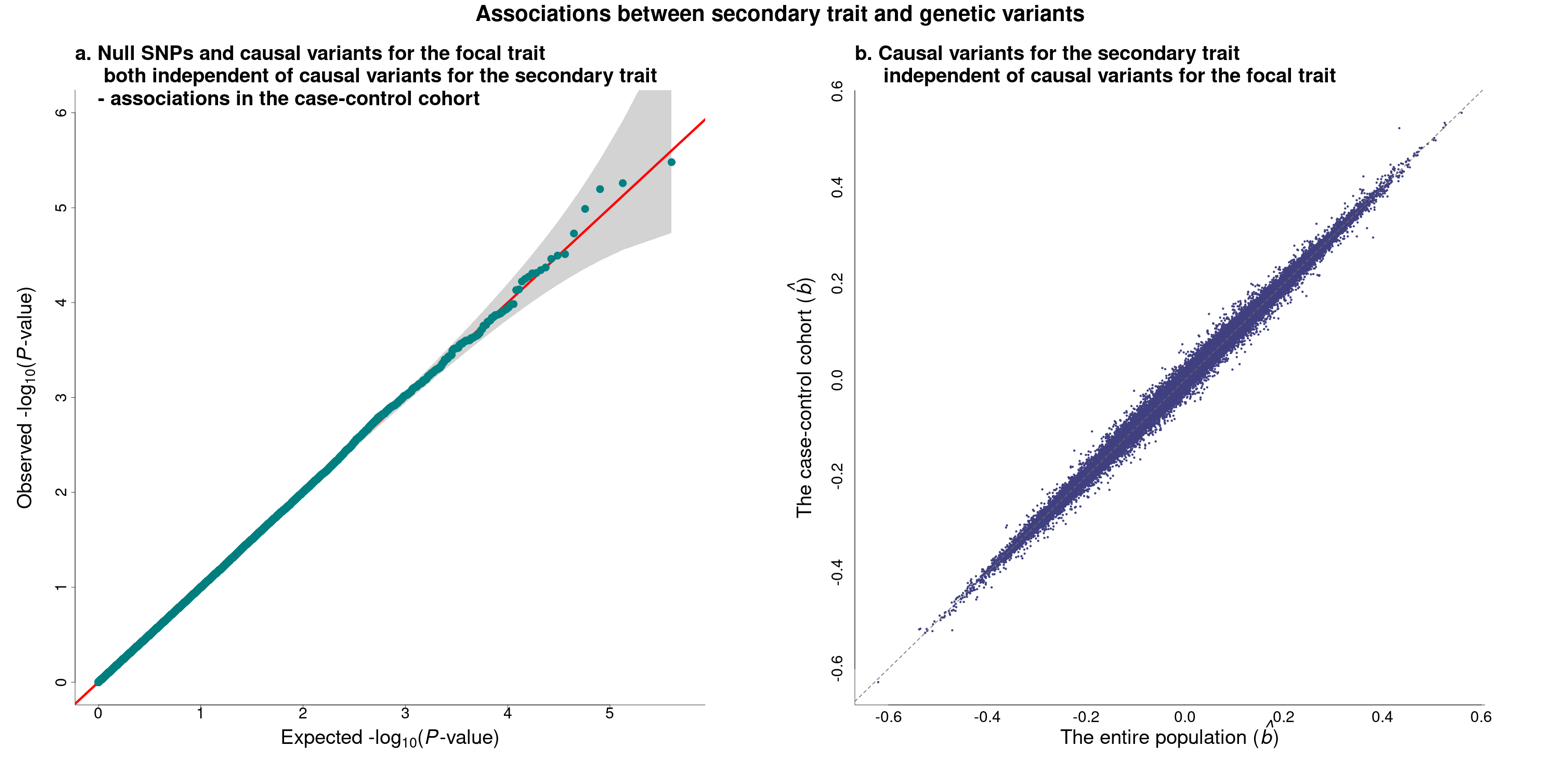

**Supplementary Figure 9** Association between secondary trait and genetic variants when the secondary trait is independent of the focal trait. The simulation method details can be found in Supplementary Methods #3. In brief, we constructed a population with 5M individuals, simulating 3 separate sets of variants without overlap in the entire population, 1) 100 causal variants for mental disorder (the focal trait, *y*), 2) 100 causal variants for the secondary trait (*x*) and 3) 100 null SNPs. Assuming the disease prevalence of *y* was 2%, there were 100K cases and 4,900K non-cases. A case-control cohort was sampled from the entire population. It consisted of two sub-cohorts, 1) 30K randomly sampled individuals, and 2) 26K randomly sampled cases. Duplicated individuals were excluded. For each simulation, we conducted the association analyses between SNP and *x* in the entire population and the case-control cohort. The simulation was conducted with 1,000 replicates. Shown in the figure were estimates of the three SNP sets, a) QQ plot of a subset of SNPs including 100 causal variants for *y* and 100 null SNPs, b) for 100 causal variants associated with *x*, comparison of effects between the entire population and the case-control cohort, and

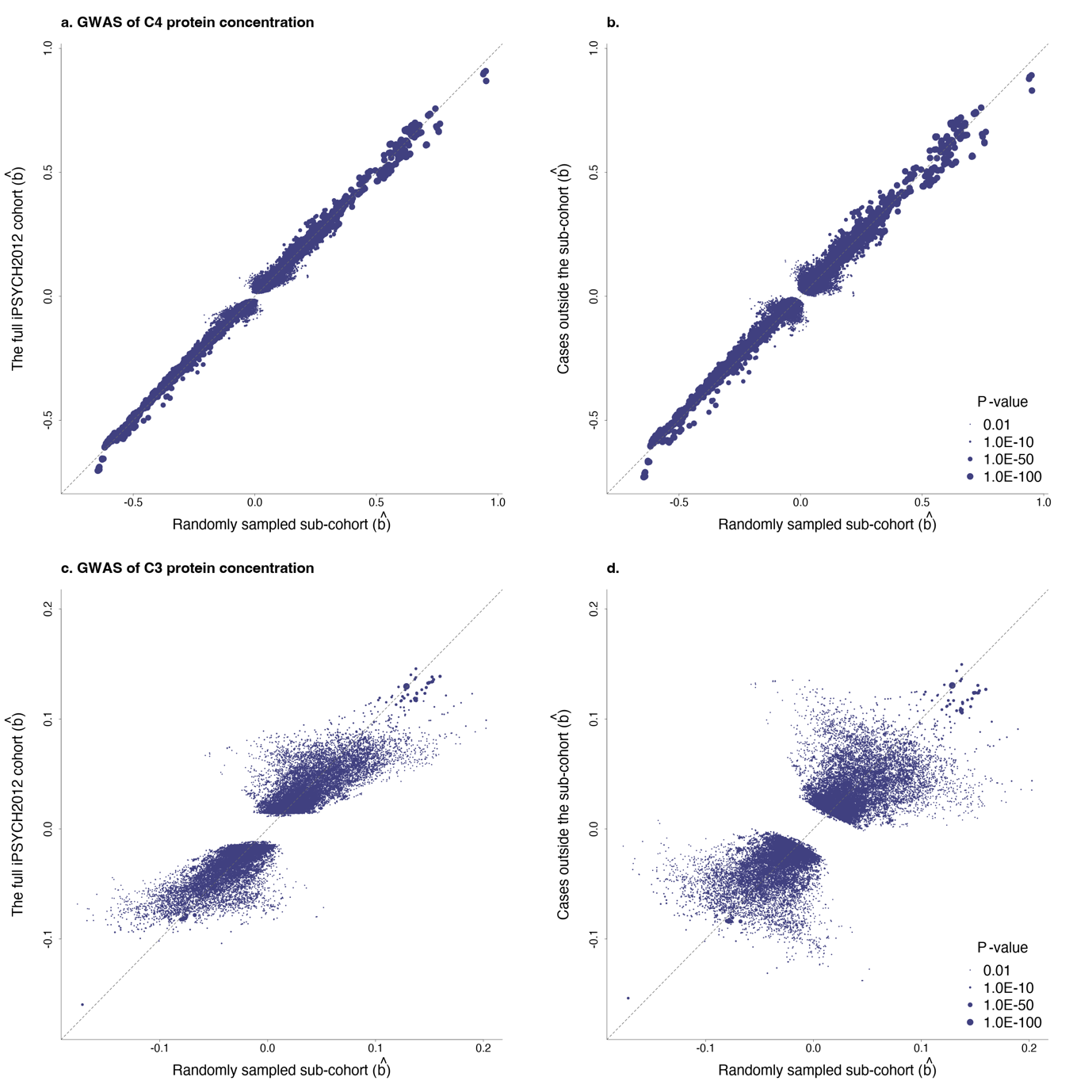

**Supplementary Figure 10** Estimates in the GWAS studies of C3 an C4 protein concentrations. The full iPSYCH2012 sample can be partitioned into two subsets, 1) the population-based sub-cohort with 25,078 randomly sampled participants and 2) the mental disorder cases (*n* = 50,686) outside the population-based sub-cohort. We examined the ascertainment bias in the GWAS studies of C3 and C4 protein concentrations by re-running the analyses in the two subsets. The top two panels showed the comparisons of estimated effects on the C4 protein concentration, (a) between the randomly sampled sub-cohort and the full iPSYCH2012 sample and (b) between the randomly sampled sub-cohort and the 6 mental disorder cases outside the randomly sampled sub-cohort. The bottom two panels showed the comparisons of estimated effects on the C3 protein concentration, (c) between the randomly sampled sub-cohort and the full iPSYCH2012 study and (d) between the randomly sampled sub-cohort and the 6 mental disorder cases outside the randomly sampled sub-cohort. For better graphic demonstration, we excluded the null SNPs (*P*-value < 0.01) in the GWAS using the full iPSYCH2012 cohort. *P*-values were truncated at 1.0E-100.

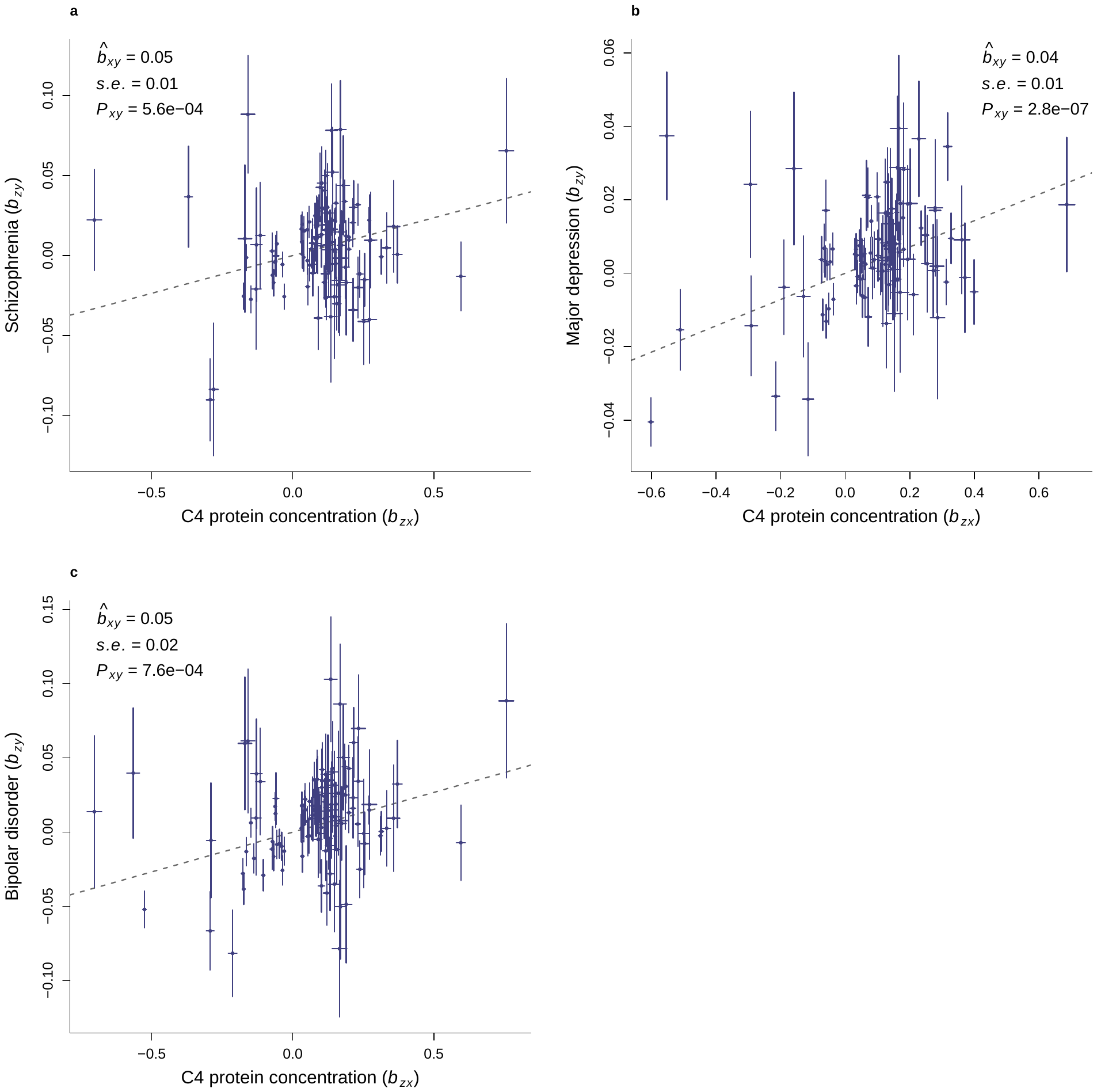

**Supplementary Figure 11** GSMR analyses of C4 protein concentration against mental disorders. The mental disorders shown in the plot were a) schizophrenia (SCZ), b) major depression (DEP), and 3) bipolar disorder (BIP). The GSMR estimates including effect, standard error and P-value were shown in each panel. Slope of dash line represented GSMR effect. Dots represented SNPs used in the analysis with potential pleiotropic SNPs excluded. Bars represented standard errors of SNPs from GWAS.

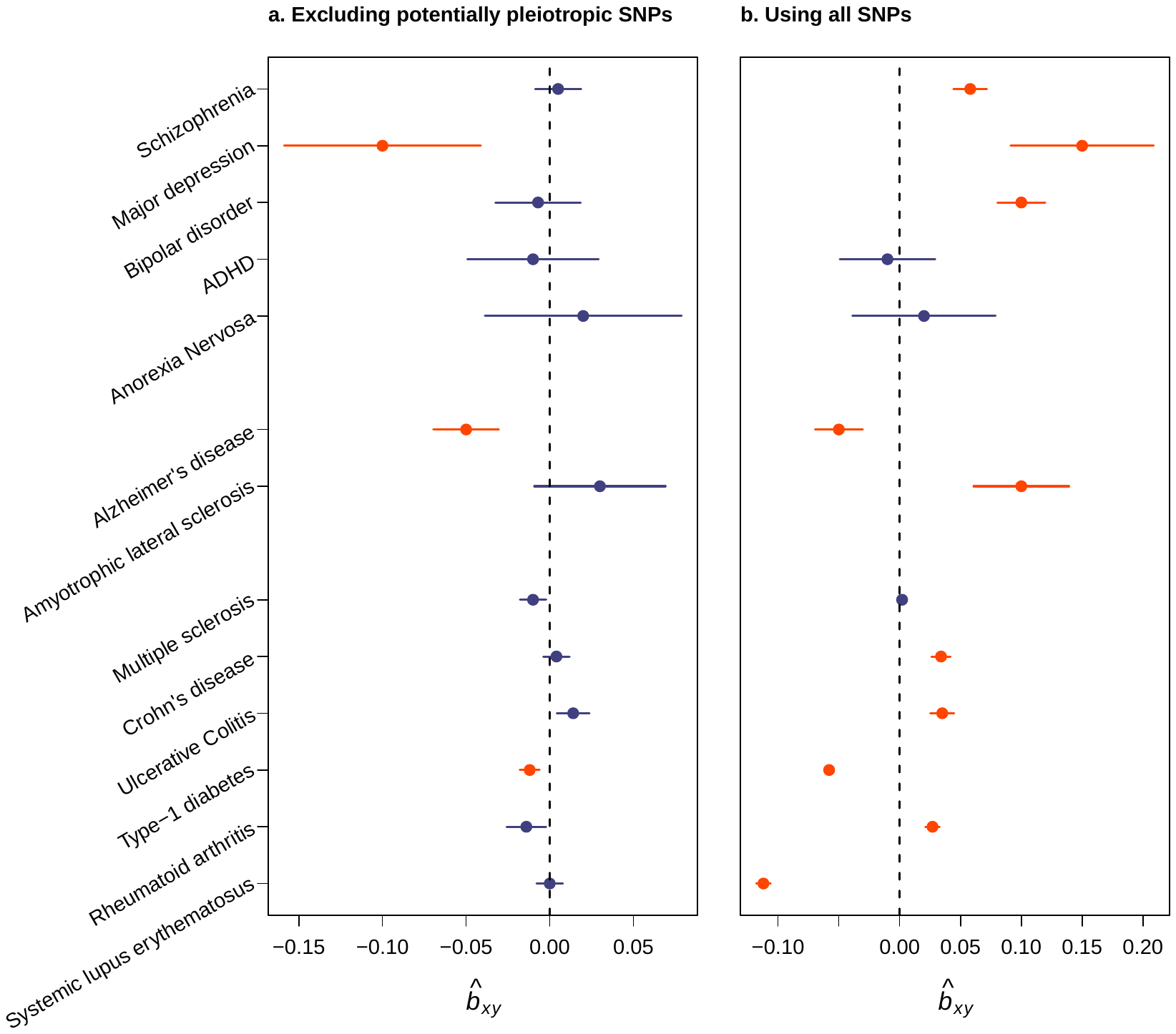

**Supplementary Figure 12** GSMR analyses to examine causation of mental and autoimmune disorders against C4 protein concentration. Shown were GSMR results from 13 mental and autoimmune disorders to C4 protein concentration. Autism was not shown in the plot, because < 5 SNP instruments were available in the GSMR analysis. The left panel (a) showed the results using the SNPs with potential pleiotropy excluded and the right panel (b) showed the results using all the SNPs. The Bonferroni corrected threshold was 1.8×10^-3^. The significant results were highlighted with red colour. Bars represented 95% confidence interval.

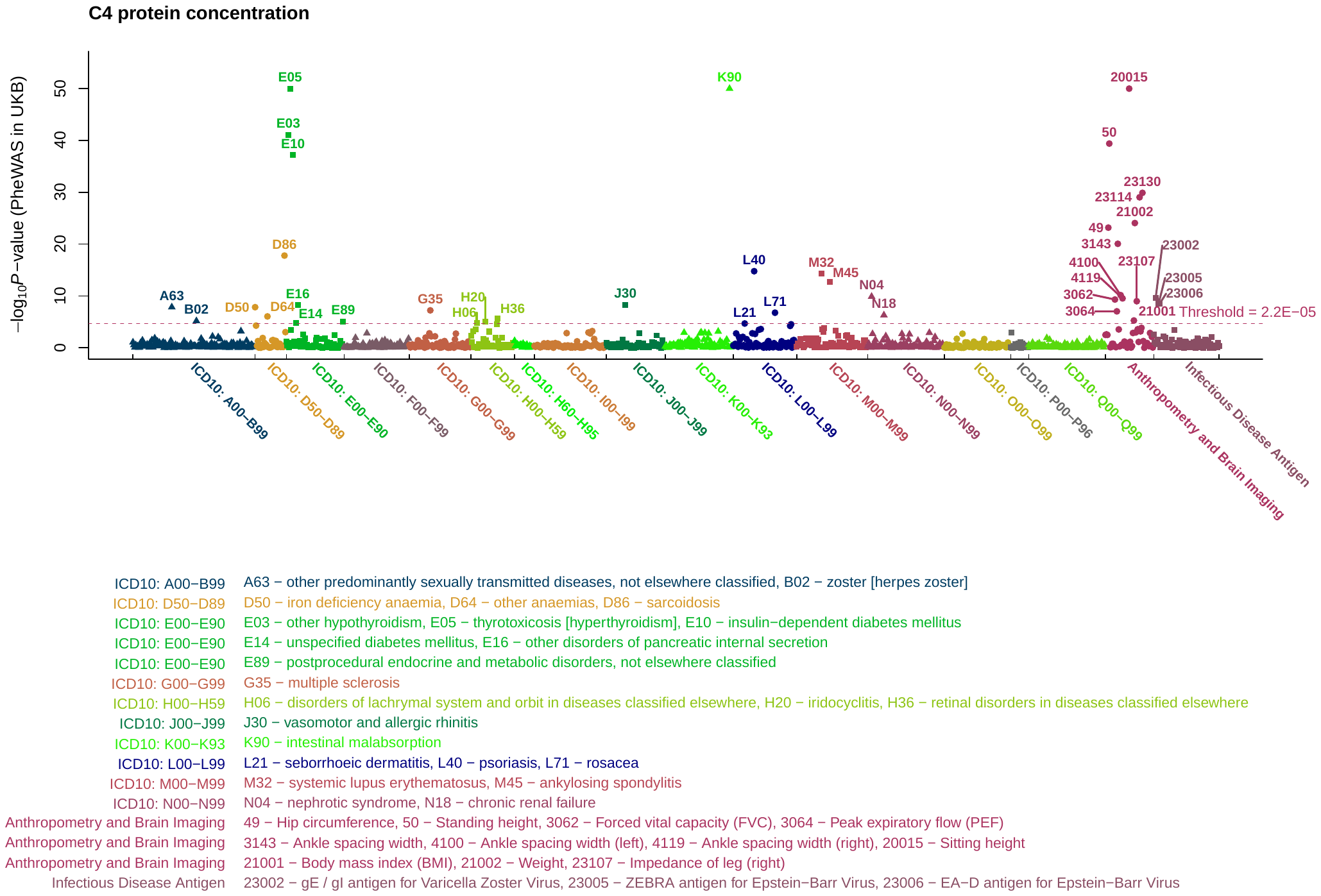

Supplementary Figure 13 Phenome-wide association study of C4 protein concentration. Shown in the figure were -log10(P-value) from the phenome-wide association study (PheWAS) between C4 protein concentration and 1,148 phenotypes in the UK Biobank (UKB) data. The polygenic risk score of neonatal C4 protein concentration was predicted by BayesR. The UKB phenotypes included 1,027 disorders, 51 anthropometric measurements and brain imaging traits, and 70 infectious disease antigens. 347,769 unrelated individuals of European ancestry were used in the PheWAS analysis. The Bonferroni corrected significance threshold was 7.3×10^-6^ (= 0.05 / (1148 × 3 × 2)). Significant ICD-10 classified disorders and the other phenotypes (anthropometry, brain imaging and antigens) were highlighted with their respective ICD-10 codes and UKB field IDs. The full names of these traits are shown below the figure.

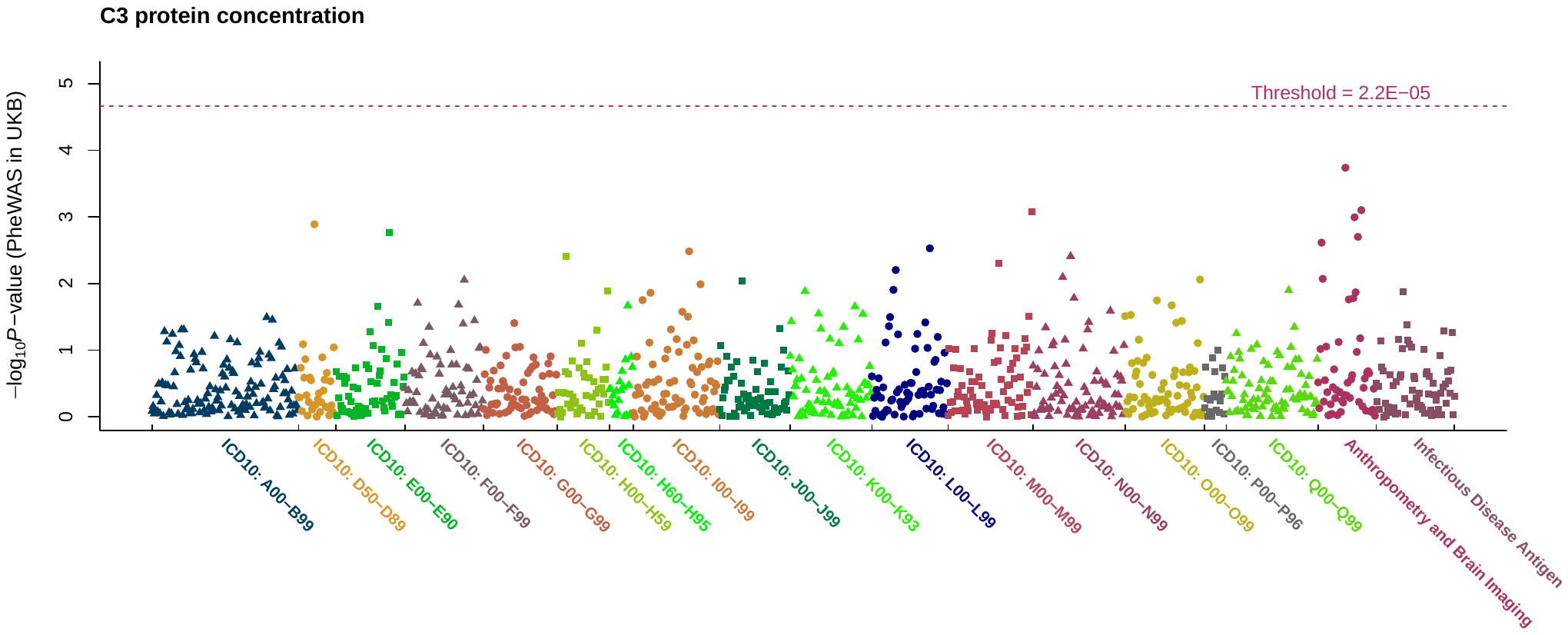

**Supplementary Figure 14** Phenome-wide association study of C3 protein concentration. Shown in the figure were -log10(P-value) from the phenome-wide association study (PheWAS) between C3 protein concentration and 1,148 phenotypes in the UK Biobank (UKB) data. The polygenic risk score of C3 protein concentration was predicted by BayesR. The UKB phenotypes included 1,027 disorders, 51 anthropometric measurements and brain imaging traits, and 70 infectious disease antigens. 347,769 unrelated individuals of European ancestry were used in the PheWAS analysis. The Bonferroni corrected significance threshold was 7.3×10^-6^ (= 0.05 / (1148 × 3 × 2)).
